# Seven replicated genomic associations of myalgic encephalomyelitis (ME/CFS): a biobank study

**DOI:** 10.64898/2026.09.09.26362115

**Authors:** Joshua Slaughter, Olivier Labayle, Breeshey Roskams-Hieter, Joshua J. Dibble, Simon J. McGrath, Sjoerd V. Beentjes, Ava Khamseh, Chris P. Ponting

**Author notes:** Equal contribution (Olivier Labayle, Breeshey Roskams-Hieter, Sjoerd V. Beentjes, Ava Khamseh). Equal contribution, alphabetical order (Sjoerd V. Beentjes, Ava Khamseh, Chris P. Ponting).

## Abstract

Myalgic Encephalomyelitis/Chronic Fatigue Syndrome (ME/CFS) is a debilitating female-biased disease with neither diagnostic biomarkers nor effective treatment nor well-understood aetiology. To investigate its biological basis, we used TarGene to perform a genome-wide association study in the UK Biobank with 1,268 ME/CFS cases, using electronic health records and survey responses to affirm ME/CFS status in cases, and non-ME/CFS status in controls. This analysis identified 176 variants as significantly associated with ME/CFS (false discovery rate < 5%). We then performed two replication studies, with similar phenotyping, in two disjoint, smaller cohorts in the UK Biobank and in the All of Us Research Program with 319 and 371 cases, respectively. Seven genomic ME/CFS risk loci replicated, although none were significant across all three cohorts. Fine-mapping at one replicated locus resolved a credible set colocalising with reduced *CLYBL* expression in putamen, in linkage disequilibrium with the replicated variant. However, the *CLYBL* Arg259 stop-gain variant was not associated with ME/CFS risk. Other replicated loci contained *BICD1*, *GRIN2A*, *CSMD1* and *RORA* genes. No gene-by-sex or gene-by-deprivation interactions survived multiple-testing correction.

**Synopsis:** 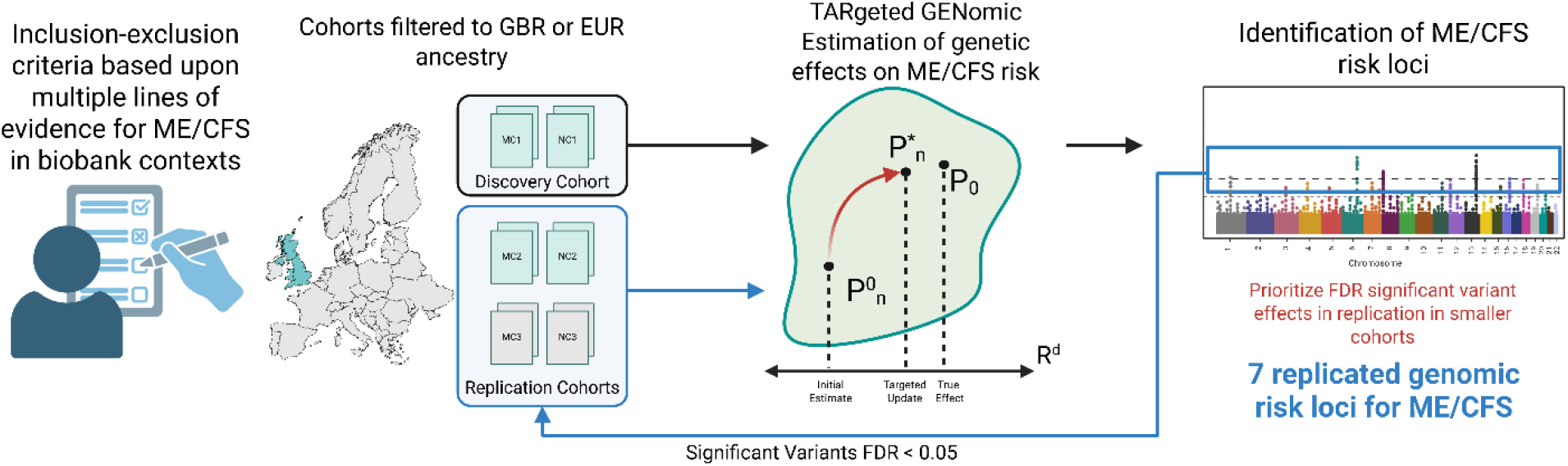

No genomic risk loci for myalgic encephalomyelitis/chronic fatigue syndrome (ME/CFS) have previously replicated across independent cohorts. We find seven variants associated with ME/CFS that replicate in disjoint biobank cohorts.

• Cases and controls are defined from multiple lines of evidence rather than a single diagnostic code.

• No variant replicates across all three cohorts, possibly reflecting differences in phenotype definition and population.

• Results provide candidate loci for follow-up into the biological mechanisms of ME/CFS.

## Introduction

Myalgic Encephalomyelitis (ME), sometimes referred to as Chronic Fatigue Syndrome (CFS), is a multi-system, female-biased illness without effective treatment, cure, or clinical biomarker. ME/CFS symptoms are heterogeneous and include post-exertional malaise (PEM), as well as persistent fatigue, muscle pain, and cognitive impairment (Committee on the Diagnostic Criteria for Myalgic Encephalomyelitis/Chronic Fatigue Syndrome *et al*, 2015). These symptoms are debilitating: those with ME/CFS experience a worse health-related quality of life than with many other illnesses (Falk Hvidberg *et al*, 2015). ME/CFS diagnosis presents substantial challenges (Bowen *et al*, 2005; Tidmore *et al*, 2015; Committee on the Diagnostic Criteria for Myalgic Encephalomyelitis/Chronic Fatigue Syndrome *et al*, 2015) despite well-established clinical diagnostic criteria for ME/CFS (Committee on the Diagnostic Criteria for Myalgic Encephalomyelitis/Chronic Fatigue Syndrome *et al*, 2015; Carruthers *et al*, 2003, 2011; National Institute for Health and Care Excellence, 2021). ME/CFS research has been hampered by these diagnostic issues and by the use of small, heterogeneous cohorts that limit reproducibility of findings (Cortes Rivera *et al*, 2019). A priority for ME/CFS research is to identify the biological factors that contribute to ME/CFS risk (Tyson *et al*, 2022), providing a foundation upon which future biomarkers and therapeutic targets might be built.

ME/CFS risk is, in part, heritable (Albright *et al*, 2011; Genetics Delivery Team *et al*, 2025). This provides the opportunity to leverage genome-wide association studies (GWAS) and extensive biobank data to elucidate gene loci whose DNA variants are associated with this risk. The UK Biobank (UKB) (Sudlow *et al*, 2015; Bycroft *et al*, 2018) and the All of Us (AoU) Research Program (Denny *et al*, 2019) provide linked phenotypic and genotypic data for their participants supporting multi-evidence case definitions and the analyses of risk-modifying factors. The findings generated from these resources have the potential to complement dedicated studies such as DecodeME which used a community recruitment strategy with clinical diagnostic criteria (Genetics Delivery Team *et al*, 2025).

Although biobanks hold multiple lines of evidence with which to affirm ME/CFS status (Samms & Ponting, 2025a; Beentjes *et al*, 2025), previous biobank studies have defined phenotypes from a single line of evidence, sometimes self-reported. In this work, we used a multi-evidence phenotyping strategy consistent with that used in a previous analysis of biobank data to uncover replicated biomarkers for ME/CFS (Beentjes *et al*, 2025). These previous studies also used conventional GWAS methods that lacked the power necessary to detect and, importantly, replicate genetic associations (Dibble *et al*, 2020; Zhou *et al*, 2018; Schlauch *et al*, 2016; Hajdarevic *et al*, 2022; Canela-Xandri *et al*, 2018). Replication across disjoint cohorts is essential to increase confidence in findings and to rule out associations due to technical and phenotypic biases.

Several factors are also suspected to modify ME/CFS risk within populations (Lacerda *et al*, 2019; Bjørklund *et al*, 2020). Genetic sex appears to be the clearest, as females are disproportionately affected over males (Bakken *et al*, 2014). Socioeconomic status (SES) has also been implicated, although findings are contradictory. Reports of ME/CFS being concentrated among high-income, highly educated women (Gunn *et al*, 1993; Samms & Ponting, 2025b; Hilland & Anthun, 2024) contrast with reports that SES does not significantly impact ME/CFS risk (Lloyd *et al*, 1990; Katz *et al*, 2009; Karfakis, 2018). This is likely to be attributed, in part, to socioeconomic barriers that hinder individuals from receiving a clinical diagnosis of ME/CFS (Samms & Ponting, 2025b). This lack of consensus highlights the need to understand further how SES may modify ME/CFS risk in populations.

To address these issues, we took advantage of state-of-the-art semi-parametric efficient and doubly-robust estimation techniques to perform our GWAS, as well as gene-by-sex and gene-by-SES analyses. These maximise power while limiting bias from model misspecification, to which conventional parametric approaches are vulnerable (Smith *et al*, 2023; Kreif *et al*, 2016). Specifically, we used TarGene (Targeted Genomic Estimation), a statistical workflow for estimating genetic effects based on the framework of Targeted Learning (van der Laan & Rubin, 2006; van der Laan & Rose, 2011; Gruber *et al*, 2024). TarGene specifically targets marginal variant effects, as well as interaction effects between genetic variants and environmental factors, and other non-linear effects (Labayle *et al*, 2025c, 2025a). TarGene’s class of estimators has been extensively applied in clinical, epidemiological, and regulatory studies (Smith *et al*, 2023; Wang *et al*, 2014; Fong *et al*, 2022; Gilbert *et al*, 2022; Lee *et al*, 2026). For the discovery GWAS, we applied TarGene to a 1,268-strong ME/CFS case cohort, together with 113,132 non-ME/CFS controls, from UKB data by applying multiple lines of evidence. To seek replication of genetic associations, we extended the phenotyping strategy to form disjoint replication cohorts within each of the UKB and AoU Research Program Biobanks. For replicated loci, we then applied fine-mapping and colocalisation with expression quantitative trait loci to prioritise candidate causal variants.

## Results

### Study populations: cases and controls

We defined three disjoint study populations for our discovery and replication case-control GWAS using criteria defined in the Methods. Cases needed to self-report a clinical diagnosis of CFS or ME/CFS at least once, a ‘fair’ or ‘poor’ overall health rating, and if they completed the Experience of Pain Questionnaire (PQ) they answered ‘Yes’ to ‘Have you ever been told by a doctor that you have Myalgic Encephalomyelitis/Chronic Fatigue Syndrome?’ Controls needed to be linked to no evidence of ME/CFS and provide a ‘good’ or ‘excellent’ overall health rating. Furthermore, participants in all observed populations must have known genetic sex being male or female, and to have predicted white British or European ancestry for the UKB and AoU biobanks, respectively. In this way, we obtained 1,268 ME/CFS cases (referred to as MC1) and 113,132 non-ME/CFS controls (referred to as NC1) for the UKB discovery cohort (UKB1). We then applied similar criteria to form two disjoint populations that we used subsequently for replication GWAS. To enable effect comparison across studies, we approximately matched the population prevalence for ME/CFS from the discovery cohort (*q*_0_ ≈ 1.10%) when forming the two replication cohorts. This value falls within a recent meta-analysis’ pooled estimate of ME/CFS prevalence, 0.89% [95% CI = (0.60%−1.33%)] (Lim *et al*, 2020; Grabowska *et al*, 2023). The first replication cohort (UKB2) contained 319 ME/CFS cases (MC2) and 28,471 controls (NC2). The second replication cohort (AoU) contained 371 ME/CFS cases (MC3) and 33,140 non-ME/CFS controls (NC3). The AoU Research Program and the UKB sampled from two distinct populations and are not known to share participants (Sudlow *et al*, 2015; Denny *et al*, 2019).

### Discovery analysis

We first performed a discovery case-control GWAS on ME/CFS risk in the UKB1 cohort. Among 670,243 genotyped (non-imputed) autosomal single nucleotide polymorphisms (SNPs) that were tested for association after quality control (Methods, <u>Quality control</u>), 176 were significantly associated with ME/CFS risk after controlling the false discovery rate (FDR) to at most 5% using the Benjamini-Hochberg procedure for multiple testing correction (Benjamini & Hochberg, 1995) (Figure 1). The summary statistics for these SNPs can be found in <u>Supplementary information</u> (S1_UKB_Discovery). We recorded a genomic inflation factor of λ = 1.09 at the median of our results (Supplementary Figure 1), indicating adequate control of population stratification (Manrai *et al*, 2019). We consider the 176 FDR-significant SNPs as prioritised candidates whose effects need to be tested for replication in independent GWAS. The replication cohorts have a smaller sample size and are therefore expected to be less well-powered relative to the discovery cohort.

**Figure 1.**
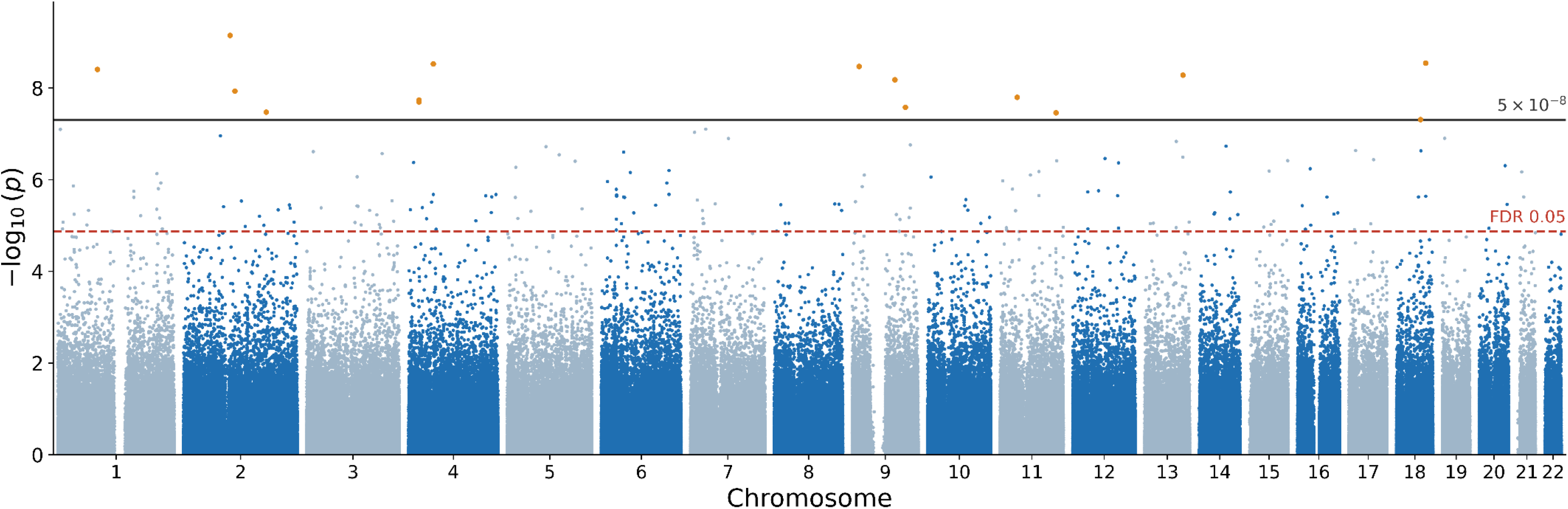
Results of the targeted estimation of the marginal genotyped variant effects in the UKB1 discovery cohort. Here, the statistical significance of the effect is shown as the -log10 of the p-value (y-axis) plotted against the variant’s chromosomal position (x-axis). The solid horizontal black line represents a frequently-applied significance threshold for genome-wide significance (−log_10_p > 7.3); the dashed red line represents the threshold when controlling the FDR at a level of α = 0.05. The results showed a modest genomic inflation at the median of λ = 1.09, indicating that confounders were appropriately controlled (Supplementary Figure 1).

Furthermore, the effects selected based on significance in the discovery cohort may be subject to the ‘winner’s curse’ and are thus expected to attenuate upon re-estimation on the smaller cohorts (Canela-Xandri *et al*, 2018; Kraft, 2008).

### Replication analyses

In the analyses of the UKB2 and AoU cohorts, we tested the set of the 176 variants associated with ME/CFS in the discovery GWAS after multiple testing correction (FDR < 0.05). Of these, eight and three variants could not be tested in UKB2 or AoU, respectively, as the minor allele count for the variants were not sufficient for stable estimation (<u>Methods, Quality control)</u>. Analysis of the UKB2 cohort replicated two significant associations (FDR < 0.05), and a further five variants were replicated in the AoU cohort (FDR < 0.05). None of these seven variants were significant after multiple testing correction (FDR < 0.05) in each of the three cohorts (Table 1, Figure 2). Further, none of 15 variants that achieved genome-wide significance in UKB1 (*p* < 5 x 10^-8^) were significant after multiple testing correction (FDR < 0.05) in UKB2 or AoU but are further discussed in Supplementary Figure 2 and Supplementary Table 1.

**Figure 2.**
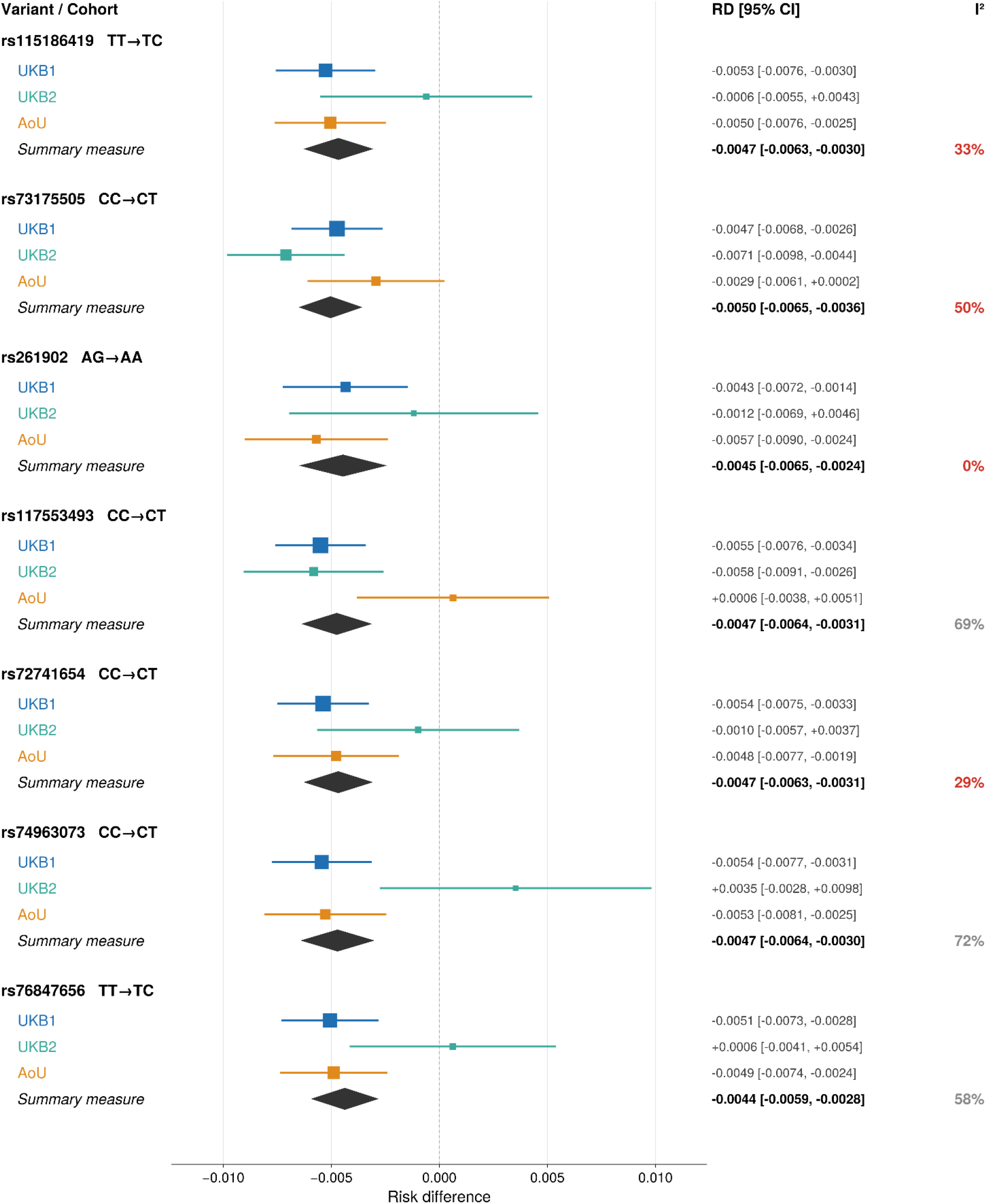
Cross-cohort effect estimates and fixed-effect meta-analysis for the seven replicated ME/CFS risk loci. For each locus, per-genotype-transition risk differences are shown for the discovery cohort (UKB1, blue), the UKB replication cohort (UKB2, green), and the AoU replication cohort (AoU, orange), with squares scaled by inverse-variance weight and horizontal bars denoting 95% confidence intervals. Here, the risk difference (RD) can be interpreted as the additive difference in the probability of ME/CFS risk between two genotypes at the same locus. The diamond, ‘Summary measure’, gives the fixed-effect inverse-variance-weighted pooled estimate across the three cohorts, and the I² statistic quantifies between-cohort heterogeneity (grey ≥ 50%, red < 50%). Effect alleles were harmonised across cohorts prior to pooling. All loci are pooled on the homozygous-major → heterozygous transition, except rs261902, where the replicated effect was estimated from the heterozygous → homozygous-minor transition. Because the discovery cohort contributes most of the inverse-variance weight and loci were selected on their prior replication, the pooled estimates summarise combined effect magnitude and cross-cohort coherence rather than providing independent confirmation of association; replication is established by the per-cohort tests (Table 1). The data were sufficiently supported in case genotype count and minor allele frequency (MAF) to estimate the risk difference at these loci: the case genotype occurred in at least 1% of the participants with ME/CFS in the observed cohort, whilst also having a MAF of at least 1% in the total population (Supplementary Figure 3, Supplementary Figure 4).

**Table 1.** Replicated variant effects across the discovery and two replication cohorts. Negative log10p values shown in bold indicate significant replicated associations with concordant effect direction. For intergenic variants, the nearest protein coding gene is shown.

| rsID | Chr:position (GRCh38) | Reference genotype | Assessed genotype | UKB1 -log<sub>10</sub>*p* | UKB2 -log<sub>10</sub>*p* | AoU -log<sub>10</sub>*p* | Nearest Gene |
|---|---|---|---|---|---|---|---|
| rs115186419 | 3:186075329 | TT | TC | **5.12** | 0.09 | **3.93** | *ETV5* (Intron) |
| rs73175505 | 8:5338479 | CC | CT | **4.93** | **6.48** | 1.15 | *CSMD1* (Intergenic) |
| rs261902 | 12:32323793 | AG | AA | **4.85** | 0.48 | **4.24** | *BICD1* (Intron) |
| rs117553493 | 13:99911262 | CC | CT | **6.53** | **3.36** | 0.11 | *CLYBL* (Intergenic) |
| rs72741654 | 15:61322139 | CC | CT | **6.19** | 0.17 | **2.90** | *RORA* (Intergenic) |
| rs74963073 | 16:10008215 | CC | CT | **5.43** | 0.57 | **3.60** | *GRIN2A* (Intron) |
| rs76847656 | 16:28261692 | TT | TC | **5.00** | 0.10 | **3.93** | *SBK1* (Intergenic) |

TarGene’s type I error control has been demonstrated using null and realistic simulations modelled on real UK Biobank data for diverse genotype frequencies and traits (Labayle *et al*, 2025c, 2025a)(<u>Methods</u>). As a sensitivity analysis, we provide results from a null simulation for the 7 replicated variants in this work, reflecting the same genotype and case-control prevalence of 1.1% in the total population of 114,400 individuals as in UKB1 (Supplementary Table 9). We demonstrate that empirical FDR is controlled at 5%, i.e., coverage of the ground truth null effect is the nominal 95%.

### Fine-mapping and colocalisation

Next, we used TarGene to test for association to ME/CFS risk within high-quality imputed regions in ±500kb windows around each of the 7 replicated variants. Estimation results of imputed variants in the 7 regions containing these loci are shown in Supplementary Figure 5**<u>-</u>**Supplementary Figure 11. We used SuSiE (Sum of Single Effects) fine-mapping with the `coloc R package` (Wallace, 2021) to obtain credible sets of likely-causal variants within these regions.

On chromosome 13, fine-mapping resolved a 95% posterior inclusion probability (PIP) credible set of five variants (Figure 3). The lead SNP at this locus, rs9585273, lies in an intron of the protein-coding gene *CLYBL*, encoding Citramalyl-CoA Lyase, whereas the other four lie 3’ of *CLYBL* (Table 2). The replicated variant at this locus, rs117553493 is also 3’ of *CLYBL* and in moderate linkage disequilibrium (LD) with the variants in the credible set (0.54 ≤ *r*^2^ ≤ 0.60).

**Figure 3.**
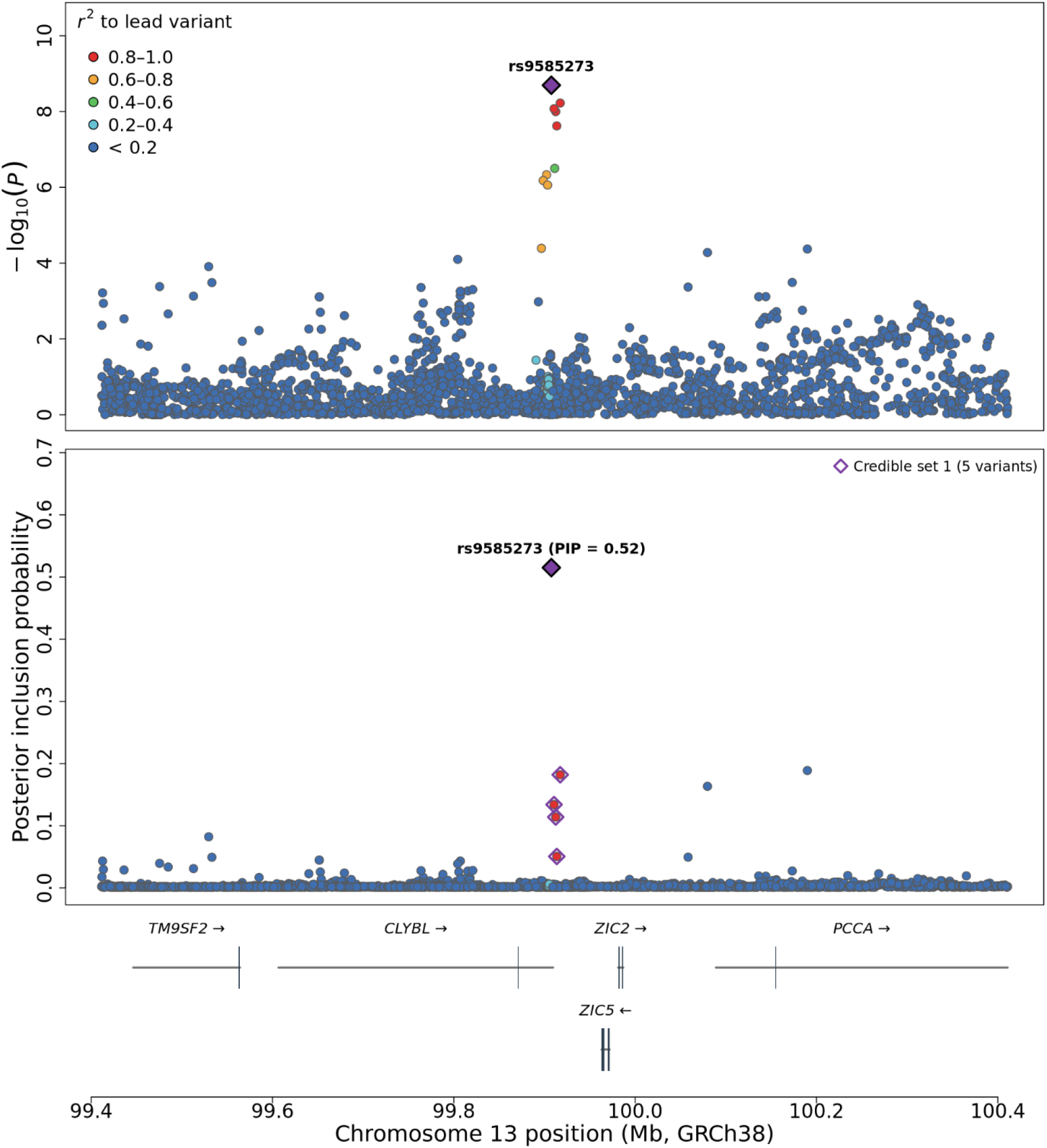
Fine-mapping analysis of the imputed set of variants in the chromosome 13 risk locus. Statistical fine-mapping of the risk locus in which the replicated association with ME/CFS risk was found at rs117553493 (in green), using the UK Biobank White-British Population for the LD reference. **(Top)** Regional associations, shown as negative log_10_(p) against chromosomal position. Points are coloured by LD (r^2^) with the lead variant (rs9585273). **(Middle)** The Posterior Inclusion Probability (PIP) for each variant indicates the probability that the specific variant is causal of the association signal. The five variants in the 95% credible set are indicated by diamonds. **(Bottom)** Protein-coding genes in the region, with exons shown as filled boxes, introns as connecting lines, and the direction of transcription indicated by arrows.

**Table 2.** Fine-mapping result of the ME/CFS risk locus located on chromosome 13. The summary statistics for the SNPs in the credible set obtained from SuSiE fine-mapping of the ME/CFS risk locus on chromosome 13 in UKB1.

| **Variant** | **Chr:position (GRCh38)** | **GWAS -log<sub>10</sub>*p*** | **Risk difference** | **Standard error** | **PIP** | **Functional consequence** |
|---|---|---|---|---|---|---|
| rs9585273 | 13:99907535 | 8.69 | -0.0054 | 9.04 × 10<sup>−4</sup> | 0.515 | 3’ UTR *CLYBL* |
| rs79477241 | 13:99917322 | 8.22 | -0.0053 | 9.03 × 10<sup>−4</sup> | 0.182 | Intergenic |
| rs9585274 | 13:99910510 | 8.07 | -0.0052 | 8.93 × 10<sup>−4</sup> | 0.134 | Intergenic |
| rs79025431 | 13:99912382 | 8.00 | -0.0051 | 8.89 × 10<sup>−4</sup> | 0.114 | Intergenic |
| rs60217909 | 13:99913530 | 7.62 | -0.0050 | 8.99 × 10<sup>−4</sup> | 0.051 | Intergenic |

One of the variants in the *CLYBL*-linked credible set, rs79025431, was significantly associated (*p* = 5.27 × 10^−9^) in the initial UKB1 discovery GWAS of genotyped variants. This SNP, although nominal (*p* = 0.0102 < 0.05), does not replicate in UKB2 after correction (FDR > 0.05), perhaps reflecting the moderate LD between variants. In summary, rs79025431 is most significantly associated with ME/CFS risk in our UKB1 discovery analysis and thus was prioritised by SuSiE fine-mapping of UKB1, whereas rs117553493 better captures the underlying signal at this locus for ME/CFS risk in the UKB2 cohort.

As 4 of these 5 variants are GTEx eQTLs for *CLYBL* in putamen (basal ganglia) tissue, we fine-mapped the same region for *CLYBL* mRNA expression using the GTEx v10 database and obtained two credible sets. Colocalisation analysis for ME/CFS risk and *CLYBL* expression using SuSiE coloc (Wallace, 2021) revealed strong support for colocalisation (PP.H4) between the credible sets for ME/CFS risk and *CLYBL* mRNA expression in putamen tissue with high posterior probability (PP.H4 = 0.994) (Table 3).

**Table 3.** Colocalisation of the chromosome 13 ME/CFS risk signal with CLYBL expression in brain putamen (GTEx v10). SuSiE fine-mapping of both traits over the 2,078 shared variants between datasets resolved a single GWAS credible set that colocalises with a CLYBL expression credible set (PP.H4 = 0.994). The four variants are common to both credible sets. GWAS and eQTL effects are aligned to the same minor allele, which is associated with lower ME/CFS risk and higher CLYBL mRNA expression; the risk allele is thus associated with reduced CLYBL expression. A second, independent CLYBL expression signal at the Arg259 stop-gain variant, rs41281112, is distinct from the ME/CFS association (PP.H3 = 0.999). RD, risk difference; PIP, posterior inclusion probability; SE, standard error.

| Signal | Variant | Position (GRCh38) | ME/CFS risk (GWAS) |  | *CLYBL* expression (eQTL) |  |
|---|---|---|---|---|---|---|
|  |  |  | RD (SE) | PIP | RD (SE) | PIP |
| **Colocalised Set**<br>PP.H4 = **0.994**, H3 = 0.006 | rs79477241 | 99,917,322 | −0.0053 (0.0009) | 0.38 | +0.55 (0.14) | 0.27 |
|  | rs9585274 | 99,910,510 | −0.0051 (0.0009) | 0.28 | +0.55 (0.14) | 0.24 |
|  | rs79025431 | 99,912,382 | −0.0051 (0.0009) | 0.24 | +0.55 (0.14) | 0.24 |
|  | rs60217909 | 99,913,530 | −0.0050 (0.0009) | 0.10 | +0.55 (0.14) | 0.24 |
| **Distinct Set**<br>PP.H3 = **0.999** | rs41281112 | 99,866,380 | — | — | −1.49 (0.23) | 1.00 |

Beyond the *CLYBL* locus, only one other locus resolved a credible set, one containing two SNPs with a PIP greater than 95% at rs73175505 on chromosome 8. However, this credible set does not colocalise with any of the queried QTL summary statistics at the locus.

### Interaction analyses

Reasoning that ME/CFS is a female-biased disease (Bakken *et al*, 2014), we used TarGene to perform a gene-by-sex interaction analysis across the set of 7 replicated variants. After correcting for multiple tests (FDR < 0.05), we did not identify significant interactions. Both rs261902 and rs76847656 have nominally significant (p < 0.05) sex-differential effects (*p* = 0.013, *p* = 0.029). For rs261902, addition of the A allele to the AG genotype increases ME/CFS risk in females with a greater effect than in males, whereas for rs76847656, addition of the C allele to the TT genotype increases ME/CFS risk in males with greater effect than in females (Supplementary Figure 12).

UKB ME/CFS participants (MC1) were associated with significantly higher area-level deprivation than controls (NC1) across the Townsend Deprivation Index (*p* = 5.3 × 10^−25^) [two-sided t-test], English Index of Multiple Deprivation (*p* = 3.2 × 10^−27^), and the Scottish Index of Multiple Deprivation (*p* = 5.4 × 10^−7^) [two-sided t-test] (Figure 4). To assess whether SED modifies genetic effects on ME/CFS risk, we again used TarGene to perform a gene-by-deprivation analysis for the set of 7 replicated variants for both measures. Again, we found no significant interactions after correcting for multiple tests (FDR < 0.05). A nominally significant interaction effect was observed at rs73175505 (p=0.024), for which addition of the risk allele T to genotype CC was increased in those experiencing higher levels of SED (Supplementary Figure 13). It is important to note that the deprivation measures were not included in the discovery or replication analyses as experienced deprivation could be a consequence of ME/CFS (Cortes Rivera *et al*, 2019).

**Figure 4.**
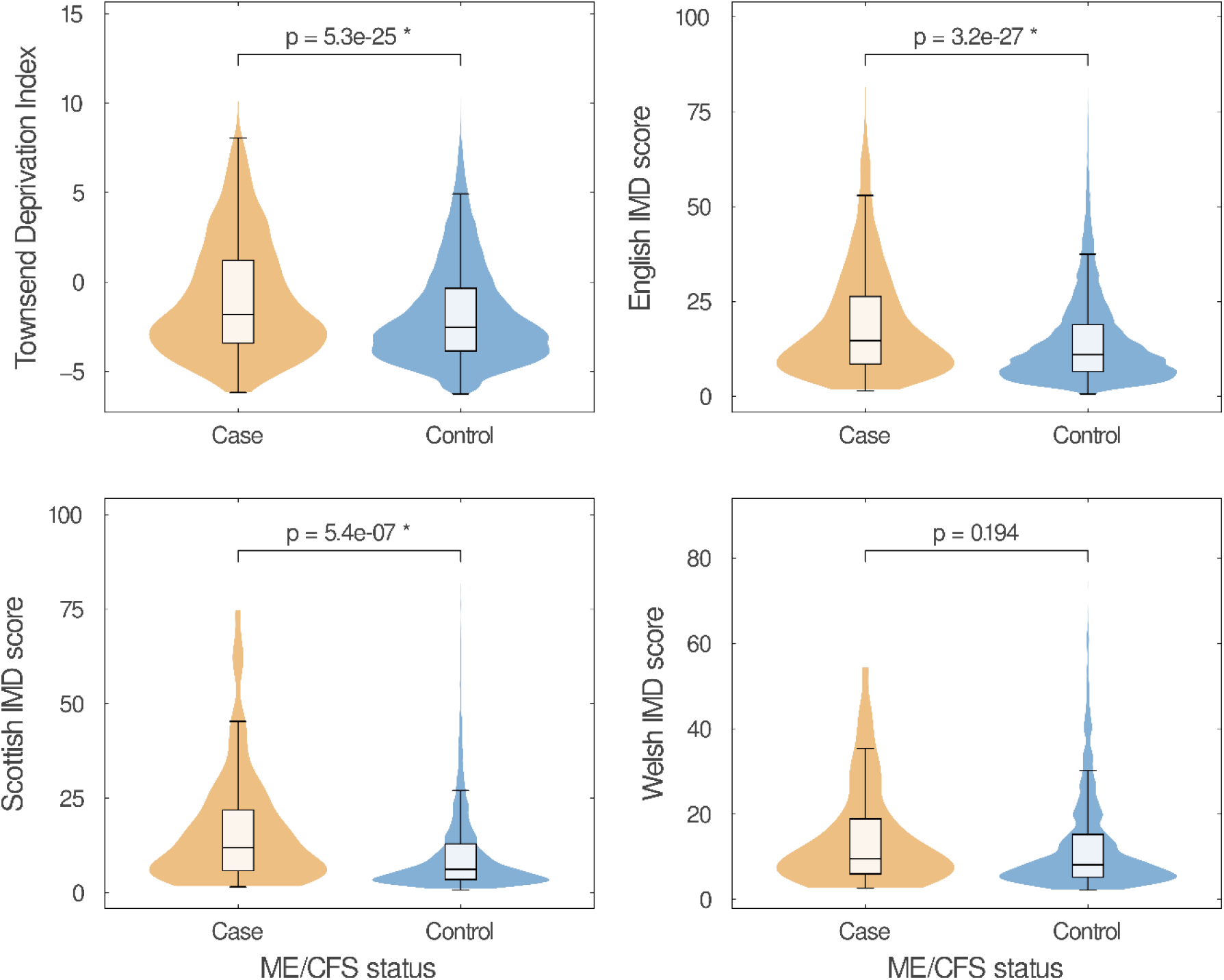
Box-and-whisker and violin plots showing the distribution of area-level deprivation for UKB1 cases and controls. (Townsend Deprivation Index [TDI]: top-left, English Indices of Multiple Deprivation [IMD]: top-right, Scottish IMD: bottom-left, Welsh IMD: bottom-right). Within the UKB1 cohort, cases generally report higher deprivation scores than those in the control group. Here, a higher score indicates that a participant resides in a more deprived area, where deprivation is characterised by the corresponding metric. An asterisk indicates that the groups are significantly different in the SED distribution. Note that because the data lacks a temporal dimension, it is not possible to conclude whether deprivation increases ME/CFS risk or whether ME/CFS status tends to result in participants being associated with socioeconomic deprivation.

### DecodeME validation

To investigate whether the replicated associations were apparent in a cohort whose cases were ascertained in a community approach using clinical ME/CFS criteria, we applied the same targeted estimation strategy as introduced in Results (<u>Discovery analysis</u> and <u>Replication analyses)</u> to 2,143 DecodeME cases with European ancestries against 191,336 White British UKB controls. Here, all UKB controls met the same criteria for NC2 as to remain disjoint from controls sampled for the discovery analysis. DecodeME cases were sampled to match the prevalence (1.1%) in the discovery analysis.

Only four of the seven replicated variants were present in the quality-controlled DecodeME genotyped data and thus could be assessed. None of the four were associated with ME/CFS risk in this analysis (Table 4). Because the prevalence was matched to that of the discovery analysis, the estimates are directly comparable with the effects from the discovery and replication analyses without rescaling. DecodeME had 83–100% power to detect effects of the magnitude estimated in discovery, yet the observed estimates were consistently close to zero. Inspection of the minor allele frequencies (MAFs) in the case and control populations at these loci further supports this lack of replication as the MAFs were differential between DecodeME cases and the cases from the cohorts used in the main analyses (Supplementary Figure 4). The associations are therefore unlikely to be present in the DecodeME cohort at the magnitude estimated in discovery, although the existence of smaller effects at these loci cannot be excluded.

**Table 4.** Look-up of replicated ME/CFS risk loci in subset of DecodeME cases; RD = Risk Difference.

| Variant | Nearest gene | DecodeME RD (95% CI) | p | Discovery RD | Power |
|---|---|---|---|---|---|
| rs115186419 | *ETV5* | -0.0014 (-0.0036, +0.0008) | 0.23 | -0.0053 | 100% |
| rs73175505 | *CSMD1* | +0.0004 (-0.0018, +0.0026) | 0.72 | -0.0047 | 99% |
| rs261902 | *BICD1* | +0.0000 (-0.0029, +0.0029) | 0.99 | -0.0043 | 83% |
| rs117553493 | *CLYBL* | -0.0002 (-0.0024, +0.0019) | 0.85 | -0.0055 | 100% |

**Table 5.** Selection criteria for three disjoint case-control cohorts that were used in discovery and replication analyses of genetic effects in ME/CFS risk. The UKB1 and UKB2 sets were filtered to predicted White British ancestry whereas the AoU cohort was filtered to predicted European ancestry. All cohorts were restricted to participants with an assigned sex-at-birth of male or female. The overlap of evidence for ME/CFS case status can be observed <u>in</u> Supplementary Figure 16<u>-</u>Supplementary Figure 18.

| **Selection Criteria** |  |
|---|---|
| **UKB1** |  |
| **Discovery: Cases (MC1), *n* = 1,268** | **Discovery Controls (NC1), *n* = 113,132** |
| ✓ Participants self-reported a CFS diagnosis in verbal interview at their first visit to a UKB Assessment Centre (UKB field 20002).<br>✓ Participants reported an overall health rating (UKB field 2178) of ‘Poor’ or ‘Fair’ at baseline.<br>✓ If the participant completed the Experience of Pain Questionnaire (PQ) (UKB field 120010) they answered ‘Yes’ to ‘Have you ever been told by a doctor that you have Myalgic Encephalomyelitis/Chronic Fatigue Syndrome?’ | ✓ Participants did not self-report a CFS diagnosis at any of the four verbal interviews at a UKB Assessment Centre (UKB field 20002).<br>✓ Participants reported an overall health rating (UKB field 2178) of ‘Good’ or ‘Excellent’.<br>✓ Participants completed the PQ (UKB field 120010) and answered ‘No’ to ‘Have you ever been told by a doctor that you have ME/CFS?’.<br>✓ Participants are not linked to a Primary Care record (CTV3 or Read v2 code) of ME/CFS.<br>✓ Participants are not linked to the ICD10:G93.3 code: 'Postviral fatigue syndrome'. |
| **UKB2** |  |
| **Replication Cases (MC2), *n* = 319** | **Replication Controls (NC2), *n* = 28,471** |
| ✓ Participants did not self-report a CFS diagnosis in verbal interviews at any visit to a UKB Assessment Centre (UKB field 20002).<br>✓ Participants reported an overall health rating (UKB field 2178) of ‘Poor’ or ‘Fair’ at baseline.<br>✓ Participants met at least one of three criteria:<br>i. Self-reporting ME/CFS through the PQ, and answering ‘No’, ‘No', and ‘Yes' to the following questions:<br>• ‘Tiredness, weariness, and fatigue goes away when resting’ (p120115, Extended Data).<br>• ‘Tiredness, weariness or fatigue happening only because of exercising and/or working too much’ (p120116, Extended data).<br>• ‘Tired after minimal physical or mental exertion’ (p120117, Extended data).<br>ii. Linkage to the read code: ICD10:G93.3 ‘Postviral fatigue syndrome’.<br>iii. Linkage to Primary Care record (CTV3 or Read v2 code) of ME/CFS. | ✓ Participants did not complete the PQ (UKB field 120010).<br>✓ Participants reported an overall health rating (UKB field 2178) of ‘Good’ or ‘Excellent’.<br>✓ Participants are not linked to a Primary Care record (CTV3 or Read v2 code) of ME/CFS.<br>✓ Participants are not linked to the ICD10:G93.3 code: 'Postviral fatigue syndrome'. |
| **AoU** |  |
| **Replication Cases (MC3), *n* = 371** | **Replication Controls (NC3), *n* = 33,140** |
| ✓ Participants had linkage to at least one of the following read codes in the AoU researcher workbench:<br>1. ICD10CM-G93.3: Postviral and related fatigue syndromes<br>2. SNOMED-51771007: Postviral fatigue syndrome<br>3. ICD10CM-G93.31: Postviral fatigue syndrome<br>4. ICD10CM-G93.39: Other post-infection and related fatigue syndromes<br>5. ICD9CM-780.71: Chronic fatigue syndrome<br>6. ICD10CM-G93.32: Myalgic encephalomyelitis/chronic fatigue syndrome<br>✓ Participants reported a rating of ‘Poor’ or ‘Fair’ to the survey question: ‘In general, would you say your health is?’. | ✓ Participants reported a rating of ‘Excellent’, ‘Very Good’, or ‘Good’ to the survey question: ‘In general, would you say your health is?’.<br>✓ Participants reported a rating of ‘Excellent’, ‘Very Good’, or ‘Good’ to the survey question: ‘In general, how would you rate your physical health?’.<br>✓ Participants reported a rating of ‘Excellent’, ‘Very Good’, or ‘Good’ to the survey question: ‘In general, how would you rate your mental health, including your mood and ability to think?’.<br>✓ Participants reported a rating of ‘Completely’ to the survey question: ‘To what extent are you able to carry out your everyday activities such as walking, climbing stairs, carrying groceries, or moving a chair?’.<br>✓ Participants reported a report of ‘None’ to the survey question: ‘In the past 7 days would you rate your fatigue on average?’.<br>✓ Participants did not report any condition code for ‘Chronic fatigue, unspecified’, ‘Chronic fatigue syndrome’, ‘Post exertional fatigue’, or ‘Postviral fatigue syndrome’. |

## Discussion

We defined ME/CFS cases and controls in biobank data by leveraging multiple lines of evidence, following Beentjes et al. (Beentjes *et al*, 2024) and Samms and Ponting (Samms & Ponting, 2025a), noting that cohorts assembled from a single line of evidence contain more participants whose recorded characteristics are discordant with ME/CFS (Samms & Ponting, 2025a). Using TarGene, we identified 176 SNPs associated with ME/CFS risk (FDR < 0.05) in the UKB1 discovery cohort, of which 15 reached genome-wide significance (*p* < 5 × 10^−8^). In the follow-up replication analysis, two associations replicated in the UKB2 cohort with a further five in the AoU cohort.

Three factors could explain why associations are not shared across all three cohorts. The cohorts were phenotyped by similar, yet different, procedures (Methods) and could reflect partially non-overlapping portions of a heterogeneous phenotype. Further, the UKB cohorts differ from the AoU cohort with respect to healthcare systems, ancestry compositions, age distributions, and health status reporting. Due to these differences, local LD structure may also vary, such that a SNP may associate with ME/CFS more strongly in one population than in another. Finally, the replication cohorts are less well-powered than the discovery cohort, and the significant effects selected on the discovery cohort are expected to attenuate upon re-estimation, in agreement with the ‘winner’s curse’.

Comparison with the initial DecodeME GWAS showed no overlap in significant loci. Further, a direct look-up of our replicated variants in DecodeME cases using the same estimation strategy found none of the replicated loci to be associated with ME/CFS risk. As control individuals overlap between the UK Biobank and DecodeME cohorts, this look-up is not formal replication. This absence of shared associations could reflect differences in case cohorts and their diagnoses. On average, DecodeME participants are about twenty years younger than UK Biobank participants. This means that more of their ME/CFS diagnoses will have been recent, and the applied criteria more often involved PEM. Further, more of their diagnoses will have been made in specialist ME/CFS services in England, which were set up from 2004. Diagnoses during referrals to some specialist ME/CFS clinics identify about twice as many alternative (i.e., non-ME/CFS) diagnoses during referrals than ordinary clinics: about 50% in specialist ME/CFS clinics (Newton *et al*, 2010; Devasahayam *et al*, 2012) versus 23% in ordinary clinics (Collin *et al*, 2012). It is likely that because ME/CFS charities were centrally involved in their recruitment, DecodeME participants were better informed about ME/CFS symptoms, especially PEM, and so had better informed GP consultations and more frequent specialist referrals. In addition, DecodeME participants are more severely affected: 71% are at least ‘moderately’ affected, meaning that they have symptoms that restrict their activities of daily living, and most have stopped work or education (Genetics Delivery Team *et al*, 2025). By contrast, all UKB participants were sufficiently well to attend a recruitment centre in person. Similarly, All of Us participants are generally older than DecodeME participants and thus are more likely to have been diagnosed using older criteria not requiring PEM. Notably, however, All of Us participants could be enrolled and provide their saliva DNA sample from home (Bick *et al*, 2024), and so may include more severely affected people with ME/CFS than UKB.

DecodeME applied clinically relevant criteria for ME/CFS, namely the Canadian Consensus Criteria (Carruthers *et al*, 2003) and the 2015 Institute of Medicine Criteria (Committee on the Diagnostic Criteria for Myalgic Encephalomyelitis/Chronic Fatigue Syndrome *et al*, 2015), which both require PEM for inclusion. By contrast, the UKB and the AoU Research Program do not capture explicit evidence for PEM. Our phenotype criteria instead relied on synthesising self-report, diagnostic codes, symptom data, pain questionnaires, and health ratings. This multi-evidence approach provided internal consistency but did not apply PEM-based criteria.

Confidence interval estimation may be sensitive to finite-sample effects in binary outcome settings with high imbalance (here case:control ∼ 1:91) combined with low heterozygote genotype frequency. In prior work with TarGene (Labayle *et al*, 2025c), using extensive null simulations, we have demonstrated the expected zero-effect statistical coverage for such scenarios. We have additionally done this for non-zero effects using realistic ancestral sampling simulations using UK Biobank data, consistently demonstrating nominal coverage of ground truth.

Here, we observe that the statistically significant variants have a preferential enrichment in major-major to major-heterozygote genotypes being associated with reduced ME/CFS probability. This is also visible directly from the raw genotype frequencies, irrespective of confidence interval estimation. As a sensitivity analysis, for the 7 replicated variants in this cohort, we added further null simulations demonstrating that if the true effect had been zero, TarGene would have offered appropriate coverage of zero, as expected (Supplementary Table 9). Nevertheless, the enrichment of significant results in one direction (protective alleles) and the possibility for additional discoveries of risk loci will need to be studied in future method development work incorporating finite-sample statistical inference techniques, applicable to complex trait genetic studies broadly.

Fine-mapping of imputed regions containing the replicated loci revealed one credible set in the chromosome 13 locus (13q32.3) with rs79477241 as lead variant. Subsequent colocalisation with GTEx v10 eQTL credible sets indicated that ME/CFS risk and *CLYBL* mRNA expression in putamen are likely influenced by the same genetic variation (PP.H4 = 0.99), with the ME/CFS risk allele associated with lower *CLYBL* expression. Colocalisation could only be investigated when fine-mapped credible sets overlapped known eQTLs. Consequently, tissue-specificity cannot be verified. The replicated variant, rs117553493, is a cis-eQTL for *CLYBL* in whole blood (Võsa *et al*, 2021). Nevertheless, the colocalised signal is only indirectly related to this replicated variant.

*CLYBL* is involved in mitochondrial vitamin B12 metabolism (Griffith *et al*, 2025), a process critical for maintaining efficient mitochondrial function and cellular energy production. Vitamin B12 deficiency or dysregulation can lead to compromised energy homeostasis, a feature often implicated in ME/CFS (Walitt *et al*, 2024). *CLYBL* belongs to a small class of polymorphic human genes for which bi-allelic variation can result in the loss of protein function. In humans, this loss-of-function (LoF) is attributable to the Arg259 truncating variant rs41281112 whose heterozygous and homozygous genotypes result in reduced or absent *CLYBL* expression, respectively (Shen *et al*, 2017; Reid *et al*, 2017; Griffith *et al*, 2025; Strittmatter *et al*, 2014; Awan *et al*, 2025). Although rs41281112 has documented metabolic effects, the LoF is not associated with a Mendelian disease phenotype, and homozygous carriers are generally healthy (Walitt *et al*, 2024). The human tolerance to partial or complete *CLYBL* LoF suggests that *CLYBL* dosage is not tightly coupled to overt physiological dysfunction. In our ME/CFS discovery GWAS, there was no evidence for association between the LoF variant and ME/CFS risk (p > 0.05), resulting in this variant not colocalising with the ME/CFS credible set in the functional analysis (PP.H3 = 0.99). In contrast, non-coding regulatory variants typically generate modest, tissue-specific differences in gene expression and mostly contribute less to risk in complex traits (Ruetz *et al*, 2019).

Six other loci were replicated in this work. rs261902 (12p11.2) lies in an intron of *BICD1* which encodes a dynein-dynactin adaptor governing neurotrophin-receptor trafficking; its biallelic LoF causes peripheral neuropathy, and common variation at this locus is associated with brain parenchymal volume in multiple sclerosis (Baranzini *et al*, 2009). rs74963073 (16p13.2) lies in an intron of *GRIN2A*, encoding the GluN2A NMDA-receptor subunit implicated in cognitive dysfunction and neuroinflammation (Hosseini *et al*, 2025). rs76847656 (16p11.2) is intergenic and resides in a gene-dense locus that also harbours *SH2B1* and *TUFM*. Additionally, rs76847656 is a cis-eQTL for *TUFM* and *CCDC101* in whole blood (Võsa *et al*, 2021). Of these, *TUFM* is implicated in mitochondrial and innate-immune functions (Liu *et al*, 2024; Li *et al*, 2026). rs73175505 (8p23.2) lies 5’ of its nearest protein-coding gene *CSMD1*, which encodes a synaptic protein that has previously been implicated in ME/CFS aetiology (Das *et al*, 2022). rs72741654 (15q22.2) lies 5’ of *RORA*, a gene implicated in inflammation pathways and that was reported at sub-genome-wide significance in a GWAS contrasting severe with less severe ME/CFS (St-Jean *et al*, 2026), although the variant reported in that study is not in LD with the replicated variant here. rs115186419 (3q27.7) lies in an intron of *ETV5,* a gene previously implicated and functionally linked to obesity (Thorleifsson *et al*, 2009; Gutierrez-Aguilar *et al*, 2014; Willer *et al*, 2009). Two of the replicated variants, at the *CLYBL* and *GRIN2A* loci, lie within ENCODE4 (Reese *et al*, 2023) candidate cis-regulatory enhancer elements. When we investigated whether genetic sex or socioeconomic deprivation modifies genetic effects on ME/CFS risk at these loci, no interaction survived correction for multiple testing, although two nominal sex-differential effects (rs261902 and rs76847656), and one deprivation-differential effect (rs73175505), were observed.

The two replication cohorts are notably smaller than the discovery cohort. Consequently, non-replication should not be interpreted as indicating the absence of an effect and better powered studies must be undertaken to further validate non-replicated associations. This does, however, mark the first ME/CFS study to report genetic associations that replicate in independent cohorts. Each of the seven replicated associations merits future experimental investigation.

## Methods

### Discovery cohort: UK Biobank

Following Beentjes et al. (Beentjes *et al*, 2024), we used three lines of evidence in the UKB to define ME/CFS cases (MC) and non-ME/CFS controls (NC) from the synthesis of primary care records, surveys, questionnaires, and verbal assessments provided by UKB participants. In the first, discovery cohort MC1, they must have self-reported a clinical diagnosis of ‘Chronic Fatigue Syndrome’ in a verbal interview at their first visit to a UKB Assessment Centre (UKB Field 20002). These cases must also have reported an overall health rating of ‘Poor’ or ‘Fair’ (UKB Field 2178) because the debilitating nature of ME/CFS likely does not justify responses of ‘Good’ or ‘Excellent’ health ratings. Furthermore, if a participant completed the Experience of Pain Questionnaire (PQ) (UKB Field 120010), cases needed to have answered ‘Yes’ to the question ‘Have you ever been told by a doctor that you have Myalgic Encephalomyelitis/Chronic Fatigue Syndrome?’.

To be considered for non-ME/CFS control cohort NC1, a participant must have completed the PQ and answered ‘No’ to the question: ‘Have you ever been told by a doctor that you have Myalgic Encephalomyelitis/Chronic Fatigue Syndrome?’ Further, participants must not have self-reported a CFS diagnosis in any of the four verbal assessment centre interviews and cannot have linkage to a ME/CFS Primary Care record (CTV3 or Read v2 code) or to the ICD10:G93.3 code, ’Postviral fatigue syndrome’.

NC1 participants must also have reported an overall health rating of ‘Good’ or ‘Excellent’ in the overall health rating survey. All participants must also have known sex-at-birth of male or female (UKB Field 22001) and were filtered by predicted ancestry of White British (see Quality control, below). Through these criteria, we identified 1,268 ME/CFS cases (MC1) and 113,132 (NC1) controls.

### Replication cohort: UK Biobank

We created a second set of UKB participants, disjoint from the first, under a separate set of definitions for cases (MC2) and controls (NC2). For MC2, we first required that an individual did not self-report CFS in the verbal recruitment interview, thereby ensuring no overlap with MC1. The overall health rating for cases was also restricted to ‘Poor’ or ‘Fair.’ To ensure that cases have ME/CFS, all in MC2 must have reported at least one of three lines of evidence: (i) self-reporting ME/CFS through the Experience of Pain Questionnaire (PQ), answering ‘No,’ ‘No,’ and ‘Yes’ to the following sequence of questions in the PQ: ‘Tiredness, weariness, and fatigue goes away when resting?’ (p120115, Extended Data), ‘Tiredness, weariness or fatigue happening only because of exercising and/or working too much?’ (p120116, Extended Data), and ‘Tired after minimal physical or mental exertion?’ (p120117, Extended Data), (ii) having a linked ICD10:G93.3 code ‘Postviral fatigue syndrome,’ and/or (iii) having a linked Primary Care record of ME/CFS.

NC2 controls did not report a diagnosis of ME/CFS upon recruitment and had neither linkage to the ICD10:G93.3 code nor a Primary Care record of ME/CFS, while reporting an overall health rating of ‘Good’ or ‘Excellent’. Like the discovery cohort, all participants must also have known sex-at-birth of male or female (UKB Field 22001) and were filtered by predicted ancestry of White British (<u>Quality control</u>). From these criteria, we defined and selected 319 UKB cases (MC2) and 28,471 controls (NC2) who overlap with neither MC1 nor NC1.

### Replication cohort: All of Us Research Program

We created a cohort of AoU participants under similar criteria as above using the All of Us Controlled Tier v8 database. For cases (MC3), a participant must have a coding positive for one of the following codes: ‘G93.3 Postviral and related fatigue syndromes’, ‘SNOMED-51771007: Postviral fatigue syndrome’, ‘ICD10CM-G93.31: Postviral fatigue syndrome’, ‘ICD10CM-G93.39: Other post-infection and related fatigue syndromes’, ‘ICD9CM-780.71: Chronic fatigue syndrome’, and ‘ICD10CM-G93.32: Myalgic encephalomyelitis/chronic fatigue syndrome’. MC3 cases must also provide a rating of ‘Poor’ or ‘Fair’ to the Overall Health Survey question ‘In general, would you say your health is?’ To be a control (NC3), a participant must not have a coding for the conditions: ‘Chronic fatigue, unspecified,’ ‘Chronic fatigue syndrome’, ‘Postviral fatigue syndrome’, or ‘Post exertional fatigue’ in their corresponding EHRs. Additionally, the NC3 participant must have provided a rating of ‘Excellent’, ‘Very Good’, or ‘Good’ to the following questions from the Overall Health Survey: ‘In general, would you say your health is?’, ‘In general, how would you rate your physical health?’, and ‘In general, how would you rate your mental health, including your mood and ability to think?’. Participants must also have provided the answers ‘Completely’ and ‘None’ to the questions ‘To what extent are you able to carry out your everyday activities such as walking, climbing stairs, carrying groceries, or moving a chair?’ and ‘In the past 7 days would you rate your fatigue on average?’, respectively. All MC3 or NC3 participants must also have known sex-at-birth of male or female and were filtered by predicted European ancestry (see Quality control, below). We identified 371 cases (MC3) and 33,140 controls (NC3) using these criteria.

### Validation cohort: DecodeME

We created a validation cohort sampling 2,143 DecodeME cases and 191,336 UKB controls, all of which met the control criteria for NC2 as to remain disjoint from controls sampled for the discovery analysis. DecodeME ascertained ME/CFS cases using clinically based criteria concordant with the Canadian Consensus Criteria (Carruthers *et al*, 2003) and the IoM 2015 Criteria (Committee on the Diagnostic Criteria for Myalgic Encephalomyelitis/Chronic Fatigue Syndrome *et al*, 2015). To place the DecodeME estimates on the same scale as the discovery analysis, cases were randomly sampled to match the discovery prevalence (∼1.1%). Of the seven replicated variants, four were present in the DecodeME genotype data following its quality control and could be assessed; the remaining three were absent and were not analysed. Because control participants are shared between DecodeME and UK Biobank, the two analyses are not independent, and this look-up therefore does not constitute formal replication.

In addition, age was not included as a covariate, as the cases and controls have non-matching age distributions (37–73 years in UK Biobank; 16–92 years in DecodeME), leaving a substantial proportion of cases without age-comparable controls. To determine whether a non-significant result reflected a true absence of association rather than insufficient power, we computed, for each testable variant, the power of the DecodeME analysis to detect the corresponding discovery effect. Power was calculated for a two-sided Wald test at α = 0.05 given the standard error observed in the DecodeME analysis.

### Defining deprivation

The involvement of SES in ME/CFS risk has been contentious in ME/CFS research as there have been several conflicting reports of its role in disease prevalence (Gunn *et al*, 1993; Lloyd *et al*, 1990; Katz *et al*, 2009; Karfakis, 2018; Hilland & Anthun, 2024). In examining the SES of UKB participants in UKB1, we used area measures for socioeconomic deprivation as an appropriate proxy. For genetic interaction analyses both Townsend Deprivation Index (UKB Field 22189) and the English Index of Multiple Deprivation (UKB Field 26410) were used as there was sufficient power. However, in examining differences in experienced deprivation levels the Scottish Index of Multiple Deprivation and the Welsh Index of Multiple Deprivation were used. Here, Townsend Deprivation Index represents a materialistic measure of socioeconomic deprivation based on an individual’s ownership and employment (Townsend, 1987). On the other hand, the Indices of Multiple Deprivation are area measures for deprivation that consider many nuanced, predefined domains such as health, crime, employment, income, living environment, education, and barriers to housing and services, specific to the context of the geography in which they are recorded (Department for Levelling Up, Housing and Communities).

In our gene-by-SED analysis, we defined deprivation using the following UKB fields: Townsend Deprivation Index (UKB Field 22189) and the English Indices of Multiple Deprivation (UKB Field 26410). These metrics are continuous variables that aim to model experienced deprivation on an output area level based upon participant postcodes. We then binarised these measures into higher-than-median TDI and EIMD scores and lower-than-median TDI and EIMD scores. This was because TarGene requires categorical variables. Furthermore, we restricted the use of the IMD to the English definition because it was the most powered and definitions across countries within the United Kingdom are nuanced and thus not comparable (Abel *et al*, 2016).

### Quality control

For the UKB1 cohort, we selected only genotyped (non-imputed) markers for analysis. These were then further filtered using PLINK2 (Chang *et al*, 2015; Shawn Purcell & Christopher Chang) to exclude those with a minor allele frequency (MAF) of less than 0.01 and those that deviated from Hardy-Weinberg equilibrium (*p* < 1 × 10^−10^) within the observed cohort. This resulted in a set of 670,243 markers for analysis in MC1/NC1. The markers selected for testing in the replication cohorts were genotyped and subjected to the same QC thresholds. In the prioritised analysis of replicated regions in UKB1, only SNPs imputed by UKB with a high-quality imputation score of 0.9 or greater were used.

Population structure was estimated via Principal Component Analysis (PCA) and the leave-one-chromosome-out (LOCO) scheme (Yang *et al*, 2014; Widmer *et al*, 2014). We performed PCA with FlashPCA2 (Abraham *et al*, 2017) on 114,400, 28,471, and 33,140 samples for each cohort, respectively (Supplementary Figure 14). From this, we obtained sets of principal components that model the population structure that might otherwise confound the relationship between genetic variant and phenotype in our analyses.

When estimating the effect of a variant on phenotype, this variant should not be included in the PCA for the adjustment set. This would otherwise cause proximal contamination that could lead to spurious associations and effect estimates (Lippert *et al*, 2011; Yang *et al*, 2013). To avoid this, we employed the LOCO scheme in which, for each tested variant, the chromosome on which the variant resides is removed from the PC estimation. This is also computationally efficient as, for a genome-wide analysis, the TarGene-GWAS pipeline would only have to compute as many sets of PCs as there are chromosomes in the analysis (e.g., 22 in an autosomal GWAS). We also excluded regions of long range, high LD (Anderson *et al*, 2010) to mitigate bias in the inference of the population structure given that these atypical regions can skew the PCA with their disproportionate contribution to the overall signal (Price *et al*, 2008).

The populations used in this study have been filtered for White British ancestry (UKB Field 22006, coding 1) or predicted European ancestry for the AoU cohort, to increase the power to detect associations and to reduce the number of PCs required for the analysis. In future studies, it is essential that more diverse populations are considered to improve the effectiveness and broader relevance of potential genomic medicine approaches for ME/CFS.

### Targeted genomic estimation

We applied workflows from the Targeted Genomic Estimation (TarGene) pipeline (Labayle *et al*, 2025c, 2026) to the case-control (MC1/NC1, MC2/NC2, and MC3/NC3) datasets to perform targeted estimation of the genetic effects on ME/CFS risk. The TarGene pipeline uses the TMLE.jl (Labayle *et al*, 2025a) Julia (Bezanson *et al*, 2012) package to support computationally efficient targeted estimation. Targeted estimation employs semi-parametric estimation theory and machine learning to estimate a causal or statistical parameter of interest from real-world data with maximal power and minimal bias (van der Laan & Rose, 2011; Gruber *et al*, 2023). This framework seeks to remove parametric modelling assumptions and to alleviate model misspecification bias whilst simultaneously accounting for population stratification. TarGene has been demonstrated to result in nominal coverage and achieve type I error control in extensive realistic simulations of the UK Biobank data (Labayle *et al*, 2025c; Labayle, 2025).

In this work we considered two statistical estimands to define our target quantities; the effect size and interaction effects. For the estimation of the genetic effect of a single variant on trait/disease (Y) as the genotype (V) changes from aa to Aa (and similarly from Aa to AA), considering population stratification W we define the Average Treatment Effect (ATE):

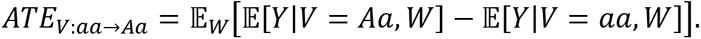

The ATE can also be interpreted as the risk difference (RD) in the context of this work.

The average interaction effect (AIE), between a variant and sex (or environmental factors) is similarly quantified as a generalisation of ATE (Labayle *et al*, 2025c; Beentjes & Khamseh, 2020) and follows as:

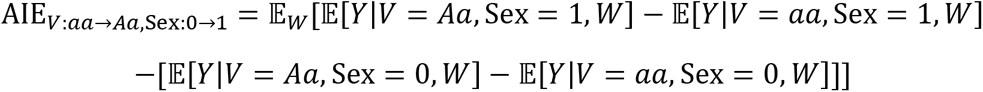

where this definition is used to quantify if the effect of a variant (aa → Aa → AA) on disease is different between males and females. It follows from the definition that if the genetic effect is not modified by sex, this interaction is zero.

We used TarGene’s GWAS workflow to estimate the targeted genetic effects on ME/CFS risk. As is common in GWAS, we selected confounders using PCs to account for population stratification, as described above (<u>Quality control</u>). Using TarGene’s PCA workflow, we identified that 6 PCs are sufficient for our populations (Supplementary Figure 14). We selected ‘Genetic sex’ (UKB Field 21022) and ‘Age at assessment’ (UKB Field 22001) as covariates associated with ME/CFS risk (Bakken *et al*, 2014).

In this study, we applied TarGene with the XGBoost algorithm (Chen & Guestrin, 2016) with hyperparameter selection via cross-validation over two key hyperparameters, maximum tree depth and L2-regularisation strength, for both the outcome regression (disease as a function of variant and PCs) and the propensity score (variant as a function of PCs) (Equation 3), because this has been shown to have optimal power and coverage in biobank-scale applications for estimating genetic effects (Labayle *et al*, 2025c; Labayle, 2025; McCaw *et al*, 2022).

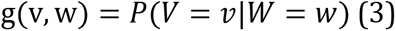

TarGene then leverages the regression and propensity score models with a semi-parametric estimator to minimise potential model-misspecification bias from these initial fits. For our semi-parametric estimator, we select weighted targeted minimum loss-based estimation (wTMLE) as it has been demonstrated to be robust in cases of near positivity violations such as estimating the unbiased effects of rare variants (Labayle, 2025; Labayle *et al*, 2025b).

To remove the assumption of a variant having a linear effect on trait, the set of genotypes at a locus is encoded as V = {*V*_0_, *V*_1_, *V*_2_}, representing the homozygous major, heterozygous, and homozygous minor genotypes respectively. The treatment effect is then estimated as the average change in outcome among the encoded genotypes. However, these tests are statistically dependent owing to the shared use of the heterozygous genotype. Therefore, joint testing is required and Hotelling’s T-squared statistic (*T*^2^) is employed to construct asymptotically valid confidence intervals and hypothesis tests (Labayle *et al*, 2025c; Hotelling, 1992). To ensure the stability of the estimates, TarGene applies a positivity constraint to each estimation task that ensures that each genotype level has a non-zero probability of occurring across all strata of confounders (Labayle *et al*, 2025c), here population stratification captured by PCs. If this constraint is violated, the query is not estimated. In our case, we applied a minor genotype frequency constraint of 0.01 following the guidance of Labayle (Labayle, 2025). In previous work (Labayle *et al*, 2025c) we have demonstrated, using null and realistic simulations modelled on UKB data via ancestral sampling, that TarGene achieves nominal coverage and control of type I error, provided the minor allele frequency is bounded from below by 0.01, see Figures 2, 3 and Supplementary Table 1 in (Labayle *et al*, 2025c). In addition, to ensure rare variants were present in both cases and controls, we implemented a filtering step that retains only those variants whose genotypes appear at a frequency greater than 1% in both case and control groups, ensuring that both groups have the alternative variant at a modest level. Finally, we applied a multiple hypothesis correction by bounding the false discovery rate (FDR) at *α* = 0.05. It is important to note that we did not have causal identifiability at the level of a variant, due to LD, meaning that associated variants may not be causal of disease risk. Rather, we identified sets of regions containing disease-associated variants in which at least one variant in each is inferred as causal of altered ME/CFS risk, strengthening the results further by fine-mapping and colocalisation analyses.

TarGene provides valid statistical inference in minimising bias due to model-misspecification and population stratification. Leveraging these favourable properties, we have obtained bias-minimised statistical estimates of genetic effects in our analyses of our discovery cohort (UKB1).

### Meta-analysis

To summarise the combined evidence at the loci that replicated in at least one disjoint cohort, we performed a fixed-effect inverse-variance-weighted (IVW) meta-analysis of the per-genotype-transition risk differences across the three cohorts (UKB1, UKB2, and AoU) (Evangelou & Ioannidis, 2013; Borenstein *et al*, 2010; Lee *et al*, 2016). For each locus, cohort-specific effect estimates were first harmonised to a common effect allele. To ensure a common estimand, each locus was pooled on a single genotype transition: the homozygous-major to heterozygous transition for all loci, except rs261902 at which there was sufficient support to estimate both transitions jointly. TarGene assesses a joint estimate by employing a Hotelling’s T² test over both genotype transitions. However, the locus was pooled on the heterozygous to homozygous-minor transition because that signal was responsible for its significance. The pooled estimate for each locus was computed as the means of the cohort-specific risk differences weighted by the inverse of their squared standard errors, with 95% confidence intervals derived from the pooled standard error.

Between-cohort heterogeneity was quantified using Cochran’s Q and the I² statistic (Higgins *et al*, 2003). We adopted a fixed-effect model because the small number of contributing cohorts (k = 3) precludes reliable estimation of the between-study variance required by a random-effects model; the reported I² instead serves to flag loci at which the cohort-specific estimates diverge. Because the discovery cohort (UKB1) contributes the majority of the inverse-variance weight, and because the meta-analysed loci were selected on the basis of their prior replication, we interpret the pooled estimates as summaries of combined effect magnitude and cross-cohort coherence, rather than as independent confirmation of association; formal replication is established by the per-cohort tests reported in Table 1.

### Gene mapping and functional annotation

We used the SNP2GENE process of the Functional Mapping and Annotation (FUMA) web application (Watanabe *et al*, 2017) to annotate our genome-wide significant ME/CFS risk loci in UKB1. We infer our sets of candidate SNPs by identifying those with nominal p-values (*p* < 0.05) and in moderate LD (*r*^2^ > 0.3) within a 1 Mb window of the lead SNP with the lowest p-value. We employed the ‘UKB release2b 10k White British’ reference panel (Bycroft *et al*, 2018) for mapping LD, due to it being the reference panel with closest genetic ancestry to the discovery cohort, UKB1.

All identified candidate SNPs were mapped to genes based on functional consequences using ANNOVAR (Wang *et al*, 2010) and the Ensembl Genes v113 database (Dyer *et al*, 2025) with FUMA. A candidate SNP was mapped to a gene if it lay within 1 kb upstream or downstream of it. When a candidate SNP was in a genomic region in which multiple genes overlapped, ANNOVAR uses prioritisation criteria to report the most deleterious function, and then only these prioritised annotations were used.

### Fine-mapping and colocalisation

We performed fine-mapping of imputed loci containing SNPs associated with ME/CFS risk in and eQTLs for such genes across several tissue contexts from the GTEx v10 database using the coloc R package (Wallace, 2021). The imputed SNPs used required an imputation score > 0.9. The `coloc.susie()` function allows for the colocalisation of multiple causal variants in the form of credible sets between two traits using SuSiE, a sparse Bayesian multiple linear regression method. For these credible sets to be identified, the LD structure of the observed variants must be known. We obtained the LD structure directly using the imputed SNPs from the UKB White British population and we approximated the LD structure for the eQTL population using the 1000 Genomes European Phase 3 data (Auton *et al*, 2015).

For colocalisation analysis, we selected the standard priors for the identification of a causal variant in ME/CFS risk: *p*_1_ = 1 × 10^−4^, identification of a causal variant in the expression of *CLYBL* in tissue *p*_2_ = 1 × 10^−4^, and the identification of a shared causal variant between the two *p*_12_ = 1 × 10^−5^. The results of the colocalisation analysis will then support one of five posterior hypotheses: here, *H*_0_ represents the null hypothesis that neither trait has a genetic association in the region, *H*_1_ represents the hypothesis that only ME/CFS risk has a genetic association in the region, *H*_2_ represents the hypothesis that only mRNA expression in tissue has a genetic association in the region, *H*_3_ is the hypothesis that both traits are associated within the region, but with different causal variants, and *H*_4_ is the hypothesis that the traits share a single causal variant (i.e. colocalisation).

## Data availability

This study used data from the *All of Us* Research Program’s Controlled Tier Dataset v8, available to authorised users on the Researcher Workbench. This research has been conducted using the UK Biobank Resource under Application Number 76173.

The datasets and computed code produced in this study are available in the following repositories:

- TarGene Bioinformatics Pipeline: GitHub (https://github.com/TARGENE/targene-pipeline)
- Semi-parametric estimation package: GitHub (https://github.com/TARGENE/TMLE.jl)
- Meta analysis, fine-mapping and colocalisation scripts: GitHub (https://github.com/edbiomedai/post_analyses_biobank_mecfs)

## Data Availability

This study used data from the All of Us Research Program's Controlled Tier Dataset v8, available to authorised users on the Researcher Workbench. This study also used data from the UK Biobank Resource under Application Number 76173, available to users on the UK Biobank Research Analysis Platform. 
The datasets and computed code produced in this study are available in the following repositories:

https://github.com/TARGENE/targene-pipeline

https://github.com/TARGENE/TMLE.jl

https://github.com/edbiomedai/post_analyses_biobank_mecfs

## Acknowledgements

This research has been conducted using the UK Biobank Resource under Application Number 76173. We thank both Gemma L. Samms and Audrey Ryback for their guidance throughout the duration of the project. This work uses data provided by patients and collected by the NHS as part of their care and support. We gratefully acknowledge the *UK Biobank, All of Us* and *DecodeME* participants for their contributions, without whom this research would not have been possible. We also thank the National Institute of Health’s *All of Us* Research Program and *UK Biobank* for making available the participant data examined in this study. CPP’s ME/CFS research was funded by NIHR and MRC (grant number MC_PC_20005). CPP, JS, and JJD were also funded by generous philanthropic donations. SB and AK acknowledge support of the UKRI AI programme, and the Engineering and Physical Sciences Research Council, for CHAI -EPSRC AI hub for Causality in Healthcare AI with real data [grant number EP/Y028856/1].

## Author information

These authors contributed equally: Olivier Labayle & Breeshey Roskams-Hieter

These authors also contributed equally: Sjoerd V. Beentjes, Ava Khamseh & Chris P. Ponting

### Authors and affiliations

Institute of Genetics and Cancer, University of Edinburgh, Edinburgh EH4 2XU, United Kingdom

Joshua Slaughter, Breeshey Roskams-Hieter, Joshua J. Dibble, Sjoerd V. Beentjes, Ava Khamseh & Chris P. Ponting

School of Informatics, University of Edinburgh, Edinburgh EH8 9AB, United Kingdom

Joshua Slaughter & Ava Khamseh

The Roslin Institute, University of Edinburgh, Midlothian EH25 9RG, United Kingdom

Olivier Labayle

School of Mathematics and Maxwell Institute for Mathematical Sciences, University of Edinburgh, Edinburgh EH9 3FD, United Kingdom

Sjoerd V. Beentjes

Health Data Research UK, 215 Euston Road, London, NW1 2BE, United Kingdom

Breeshey Roskams-Hieter

Center for Targeted Machine Learning and Causal Inference (CTML), School of Public Health, University of California, Berkeley, 94720, United States of America

Sjoerd V. Beentjes & Ava Khamseh

### Contributions

**JS:** Formal analysis; Software; Investigation; Data curation; Writing - Original Draft; Writing

- Review and editing; Visualization. **OL:** Formal analysis; Software; Investigation. **BRH:** Software; Investigation. **JJD:** Data curation; Investigation. **SJM:** Investigation. **SVB:** Data Curation; Investigation; Methodology; Formal Analysis; Software; Project administration; Writing - Original draft; Writing - Review and editing. **AK:** Conceptualization; Investigation; Methodology; Formal Analysis; Software; Supervision; Writing - Original draft; Project administration; Writing - Review and editing. **CPP:** Conceptualization; Supervision; Writing

— Original draft; Project administration; Writing - Review and editing.

### Corresponding authors

Correspondence to Sjoerd Beentjes, Ava Khamseh, and Chris Ponting.

---

**Supplementary Figure 1.**
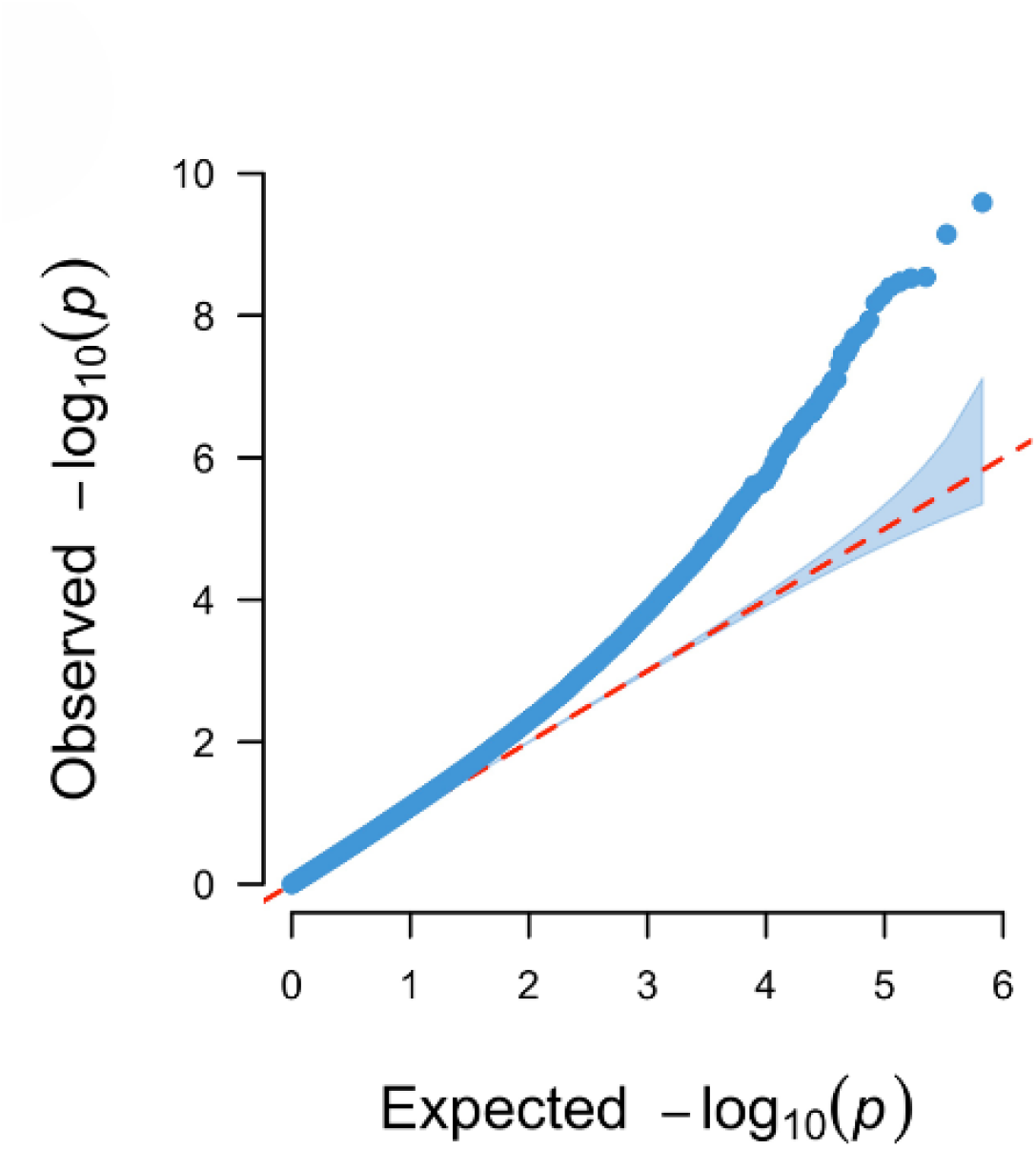
Q**u**antile**-quantile plot (QQ plot) for the UKB1 discovery GWAS.** This plot displays the departure of our genetic effect size estimates from the null uniform distribution with a modest genomic inflation at the median of λ = 1.09.

**Supplementary Figure 2.**
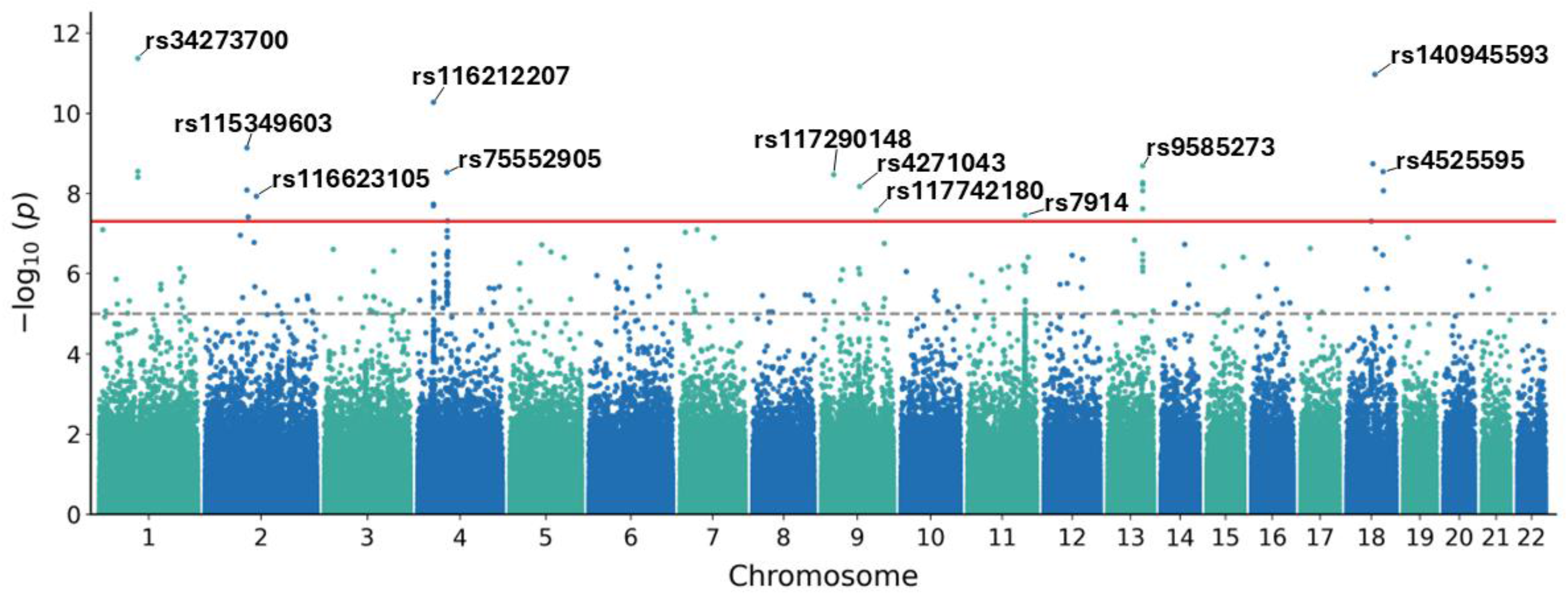
A Manhattan plot in which the imputed regions around genome-wide significant genotyped SNPs from the initial discovery analysis were tested. Here, the line (red) represents genome-wide significance (−log_10_p > 7.3) whereas the line (grey-dotted) represents the FDR level used to determine significant associations to test for replication in the disjoint cohorts. The 12 loci defined by FUMA are labelled by their lead variant.

**Supplementary Figure 3.**
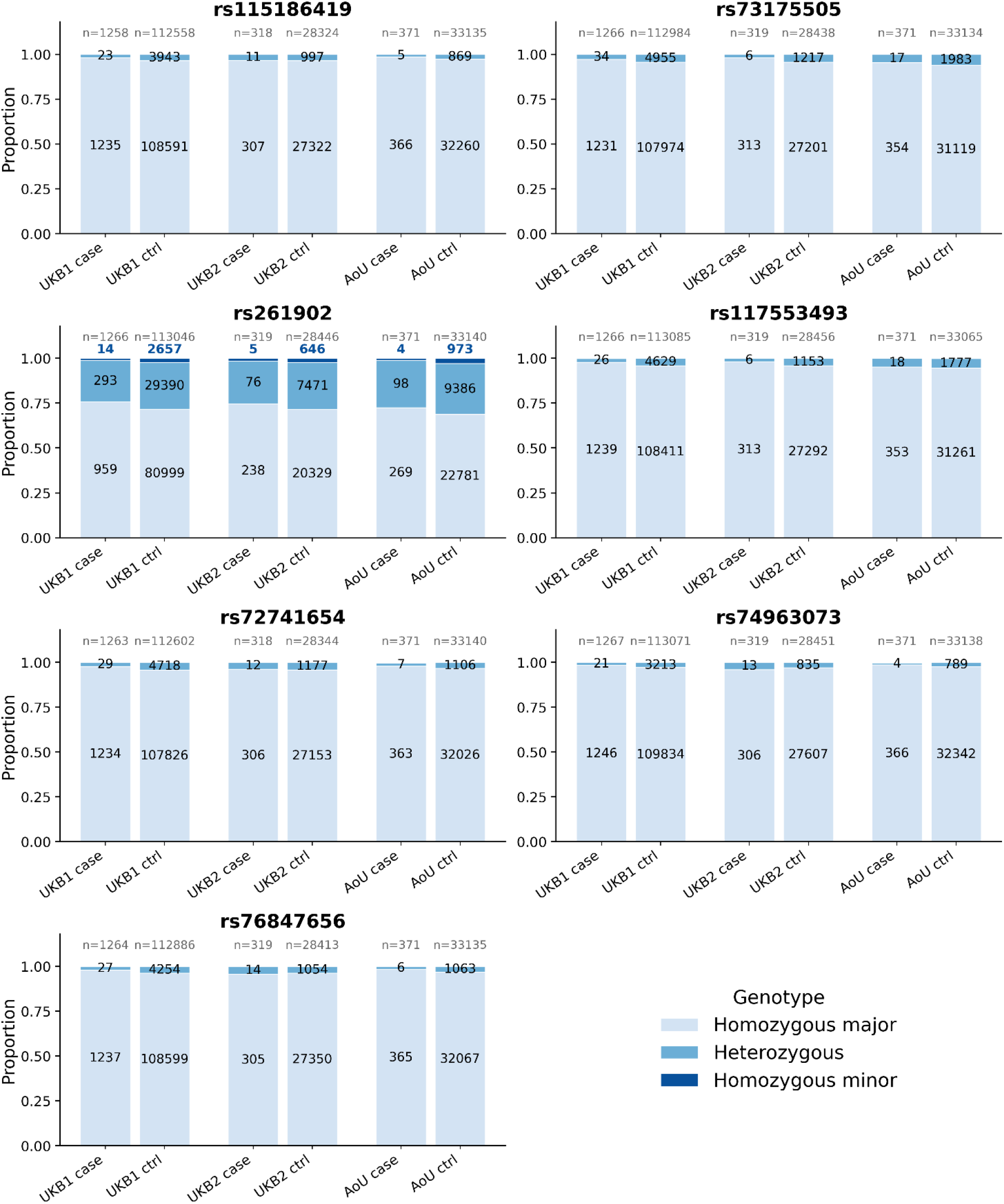
T**h**e **genotype counts across all replicated loci between the discovery analysis and at least one of the replication analyses.** Here, for each estimated effect at these loci, the count of the assessed genotype never falls below 1% of the total case count. This threshold is used as a precautionary measure to ensure that we have support across cases in *finite states even when there are no positivity violations due to a much larger control population*.

**Supplementary Figure 4.**
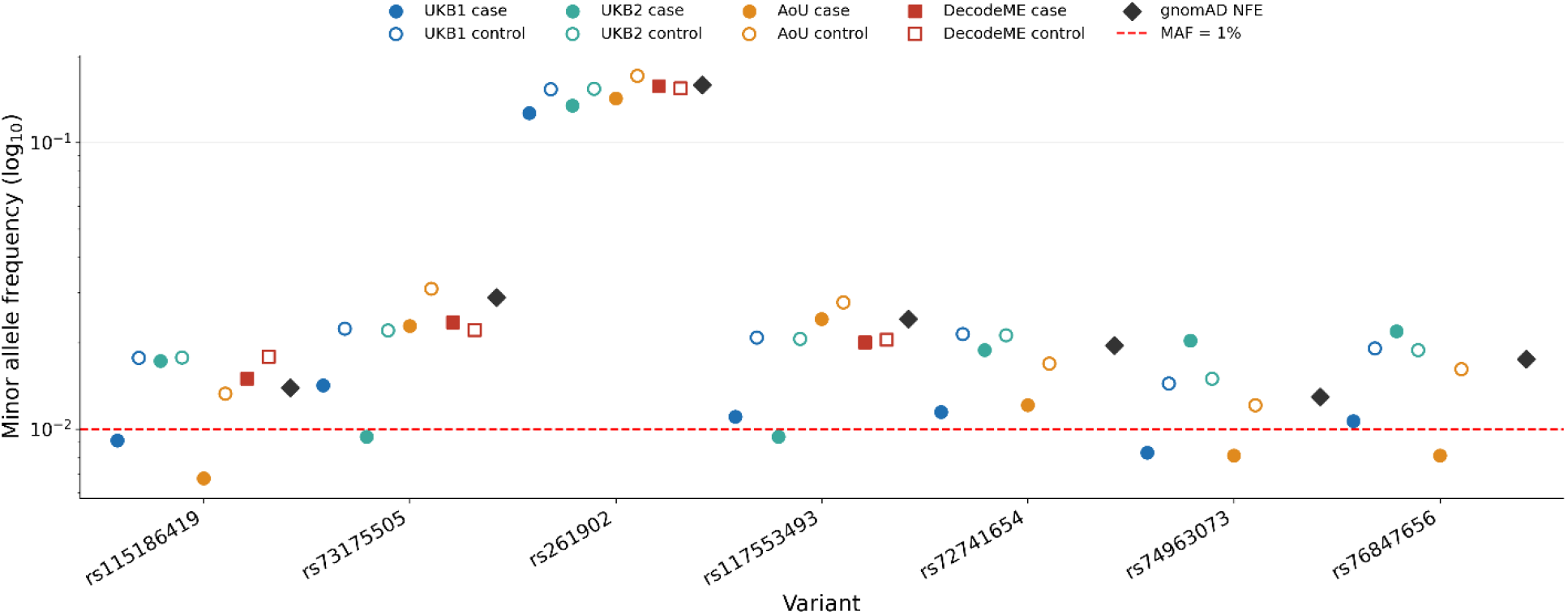
Minor allele frequencies (MAFs) at each of the replicated loci (where they could be evaluated) compared with the gnomAD (Karczewski et al, 2020) non-Finnish European MAFs for reference. Only four variant MAFs could be calculated from the DecodeME cohort as three of them were removed from the genotyped set after quality control.

**Supplementary Figure 5.**
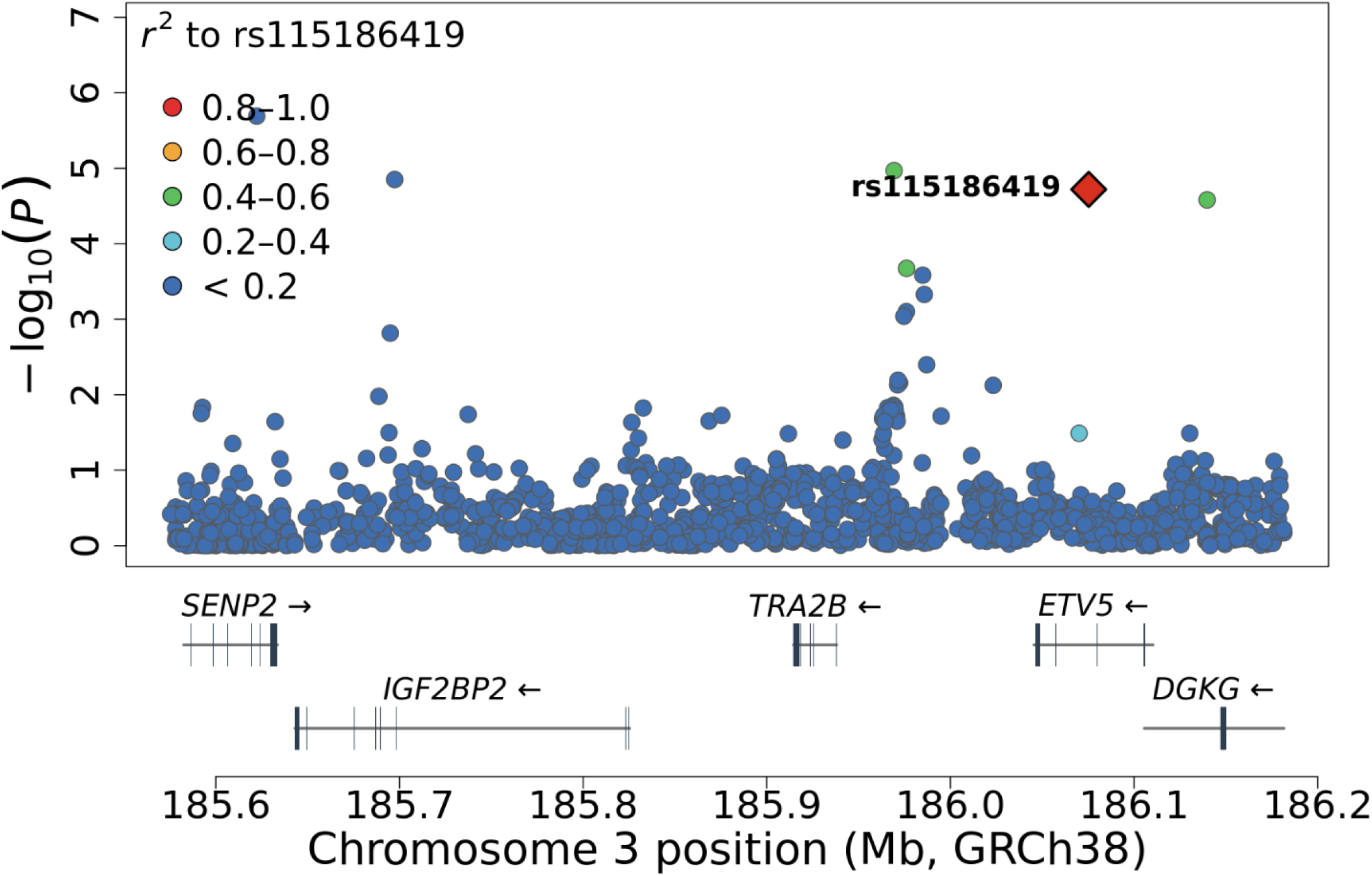
R**e**sults **from the targeted analysis of imputed regions containing the replicated variant, rs115186419.** This variant is found to be in an intron of ETV5 with moderate LD structure spanning intergenic regions and the nearby gene DGKG.

**Supplementary Figure 6.**
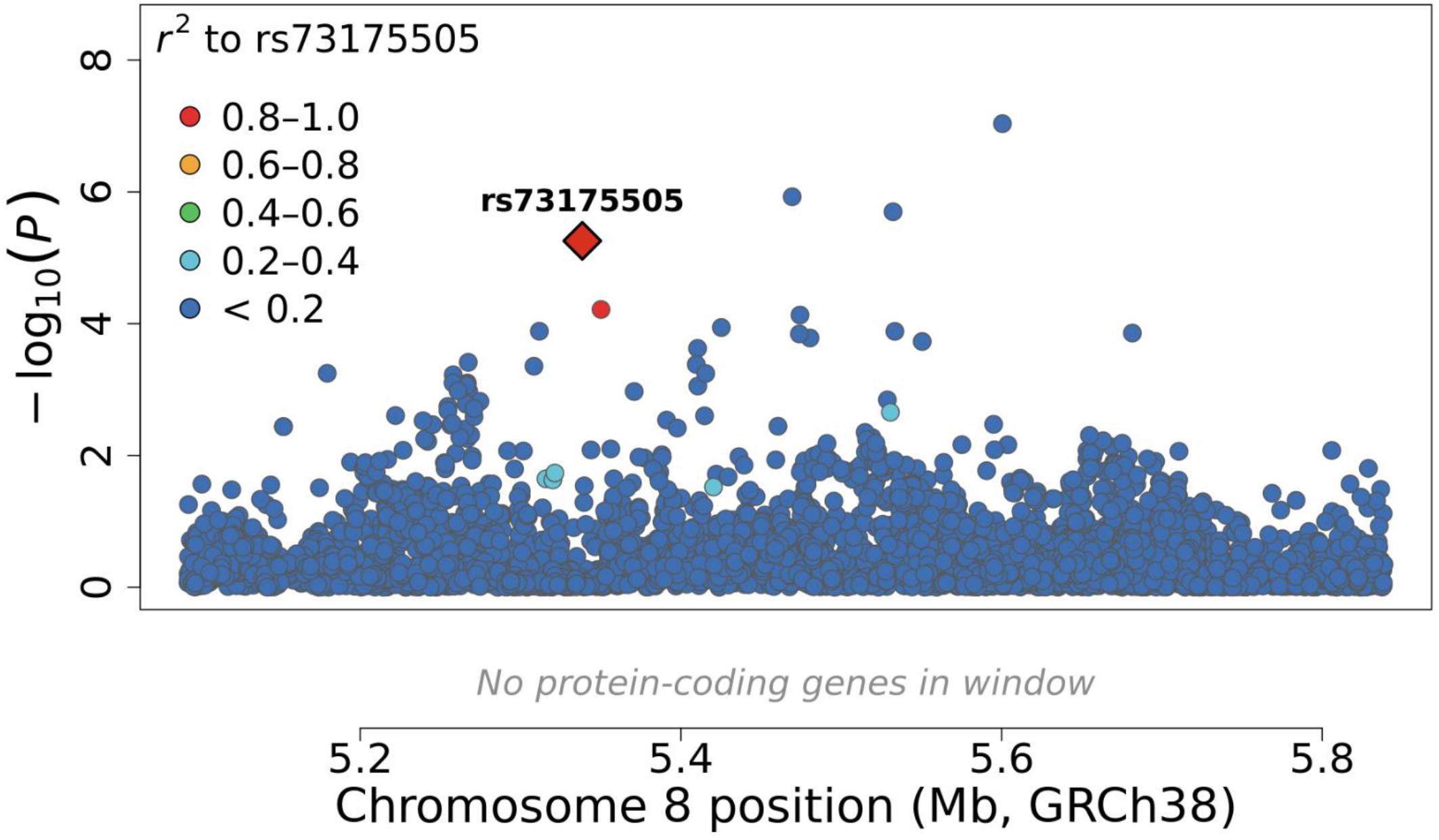
R**e**sults **from the targeted analysis of imputed regions containing the replicated variant, rs73175505.** This variant lies in a gene sparse region with no known linkage to any protein coding gene.

**Supplementary Figure 7.**
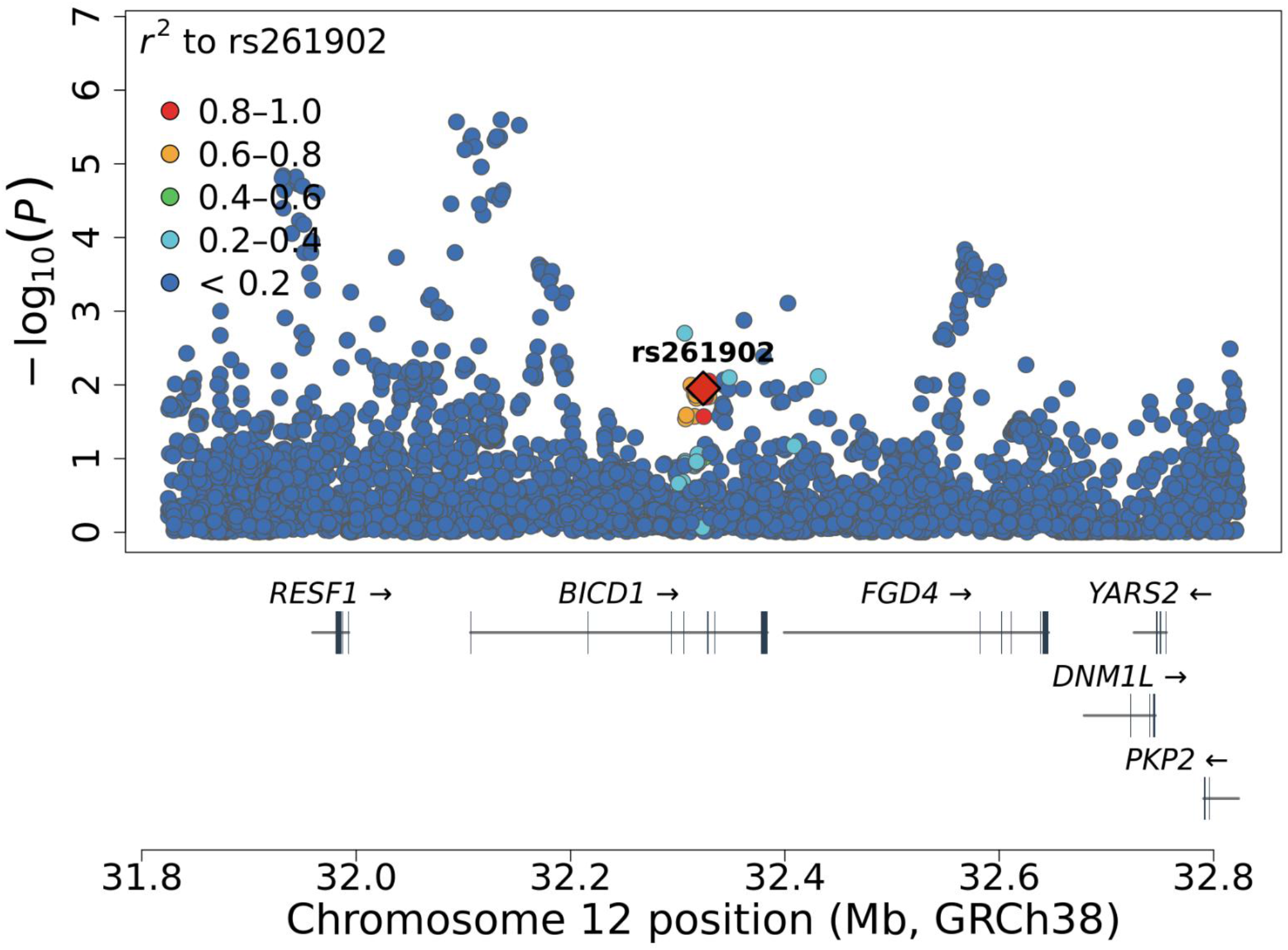
R**e**sults **from the targeted analysis of imputed regions containing the replicated variant, rs260192.** This variant is in an intron for BICD1 and shows linkage with variants spanning the nearby FGD4 gene. Note that there are many variants that are more significantly associated with ME/CFS risk in the discovery cohort than the replicated variant, rs261902 (indicated by a diamond). Some of these were among the genotyped set of 176 significant (FDR<5%) variants associated in the discovery GWAS.

**Supplementary Figure 8.**
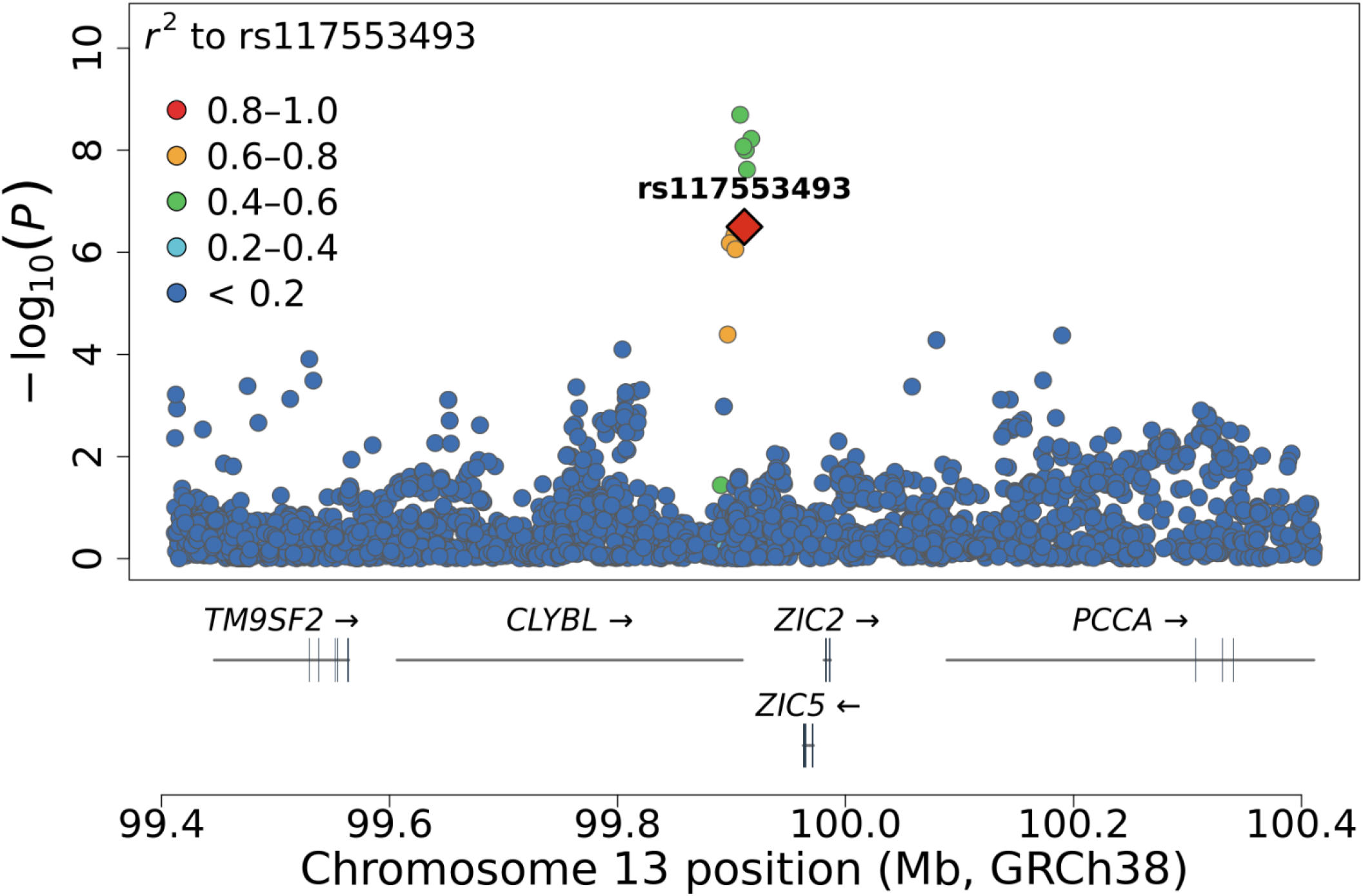
R**e**sults **from the targeted analysis of imputed regions containing the replicated variant, rs117553493.** This variant lies in an intergenic region 3’ of CLYBL and is in moderate linkage with variants that are within introns of CLYBL.

**Supplementary Figure 9.**
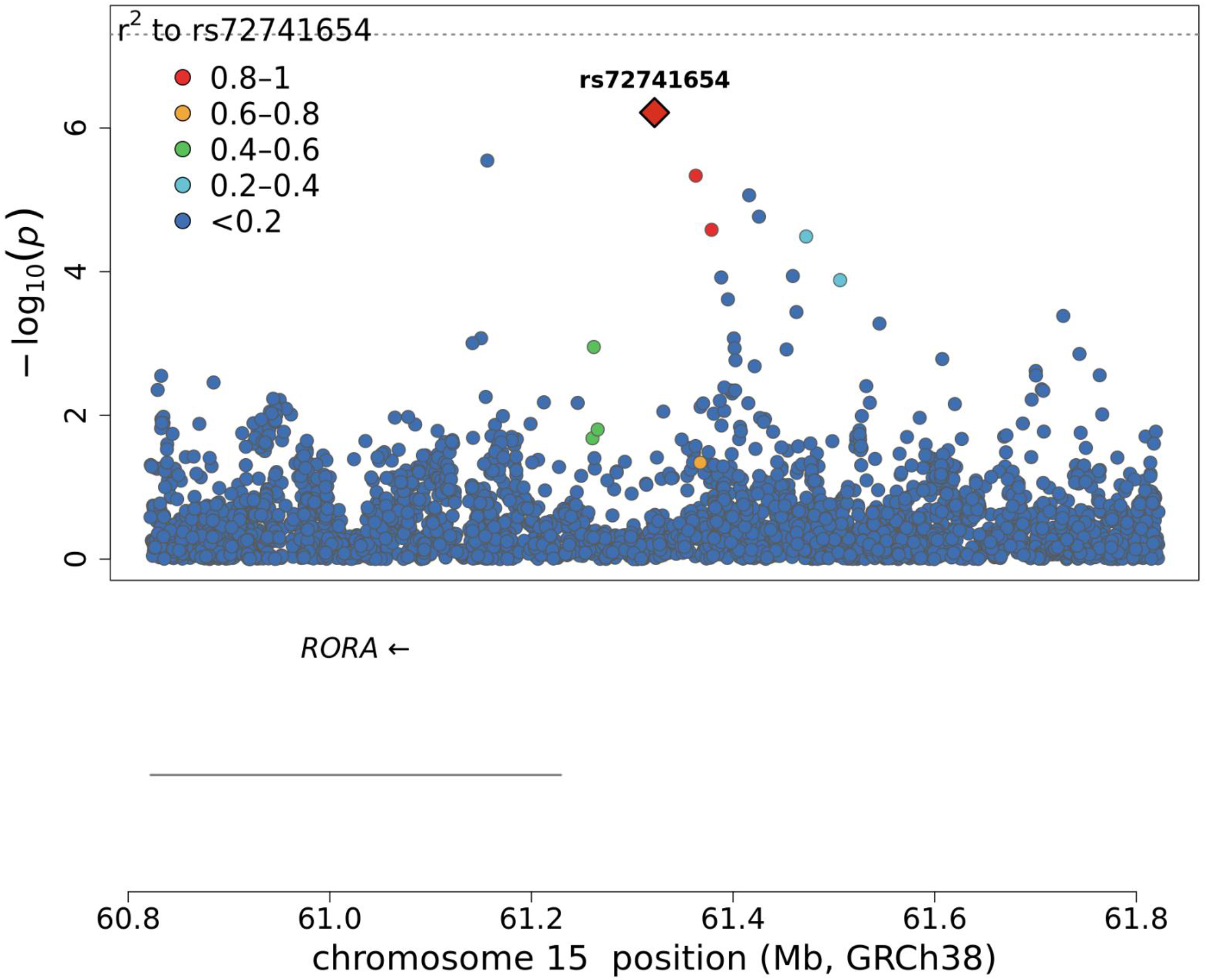
R**e**sults **from the targeted analysis of imputed regions containing the replicated variant, rs72741654.** This variant lies in an intergenic region on chromosome 15 near the RORA gene.

**Supplementary Figure 10.**
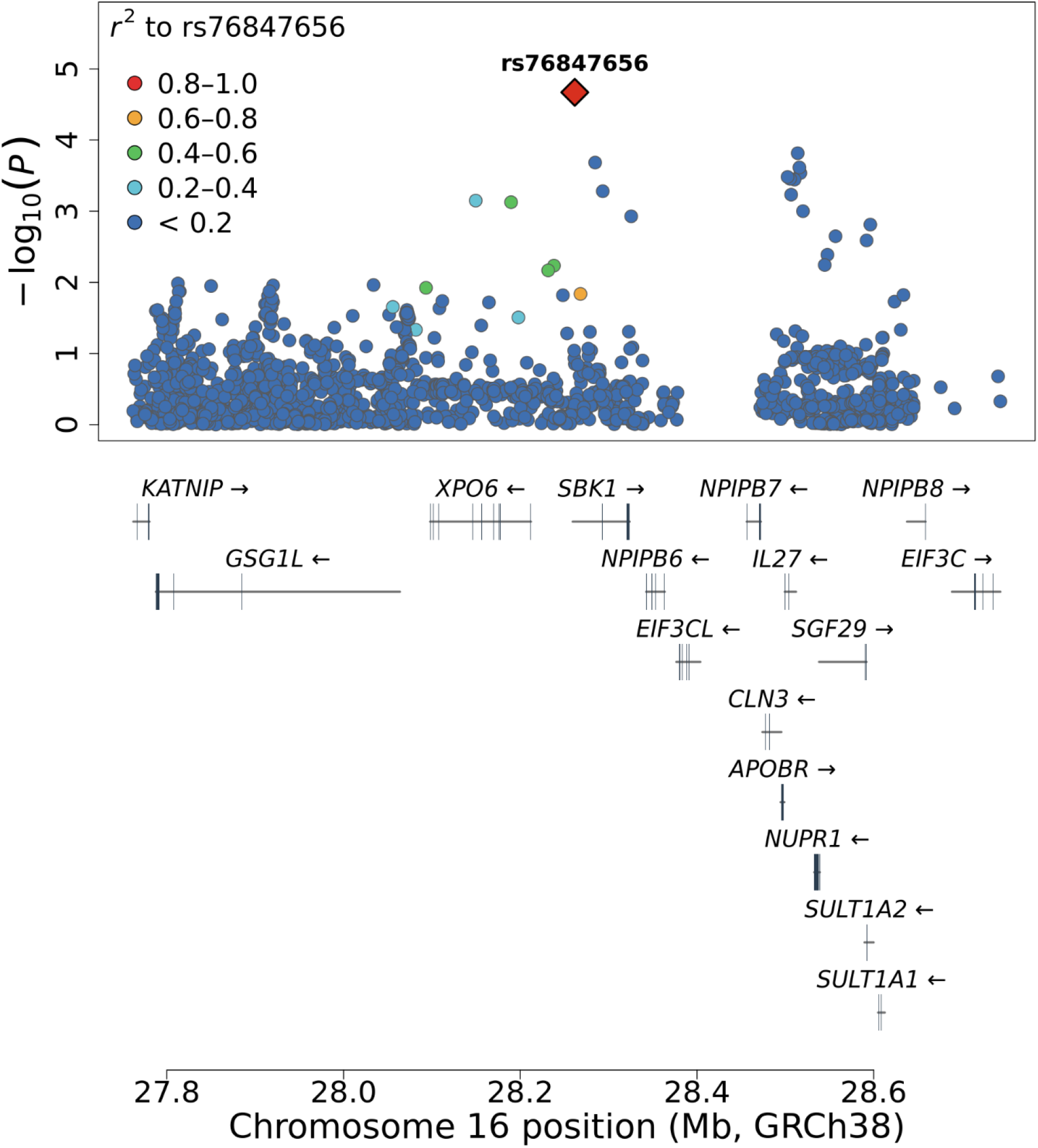
R**e**sults **from the targeted analysis of imputed regions containing the replicated variant, rs76847656.** This variant lies in an intergenic region of chromosome 16 and is in linkage with nearby variants that are within introns of XP06 and SBK1.

**Supplementary Figure 11.**
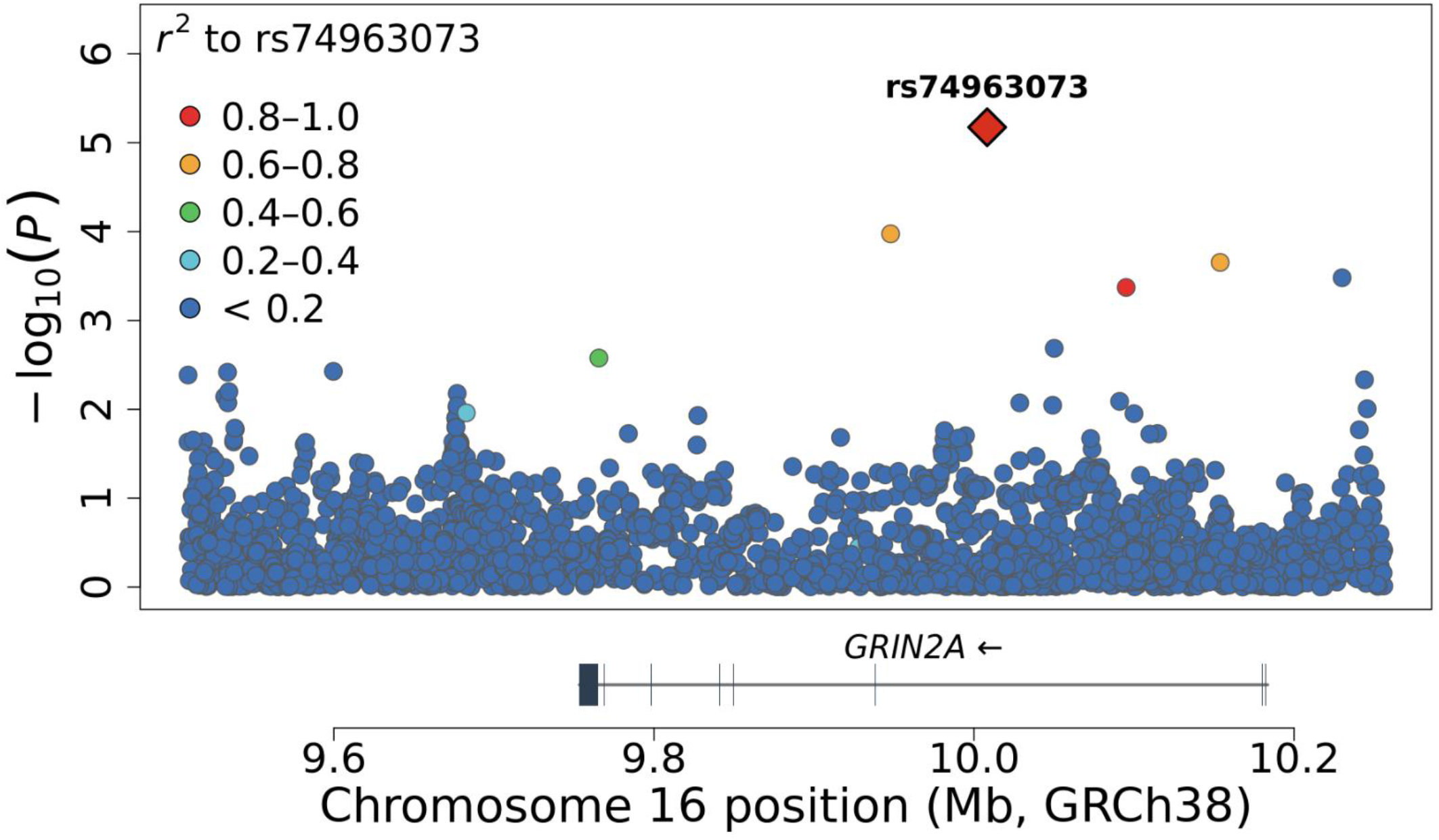
R**e**sults **from the targeted analysis of imputed regions containing the replicated variant, rs74963073.** This variant and variants in moderate linkage disequilibrium lie within introns of GRIN2A.

**Supplementary Figure 12.**
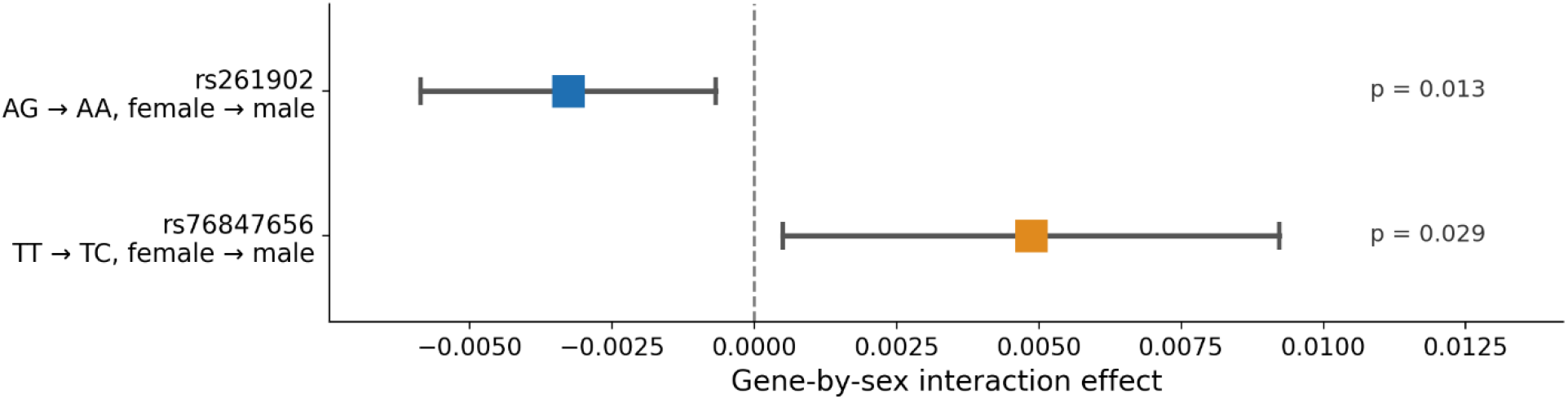
A**v**erage **interaction effects (AIE) of genotype-by-sex on ME/CFS risk for rs261902 (AG→AA) and rs76847656 (TT→TC).** Each point is the interaction effect on the risk-difference scale for the female → male sex contrast (based upon genetic sex). A negative value indicates a larger genotype effect in females and a positive value a larger effect in males. Bars show 95% confidence intervals. Both interactions are nominally significant but do not survive correction for multiple testing.

**Supplementary Figure 13.**
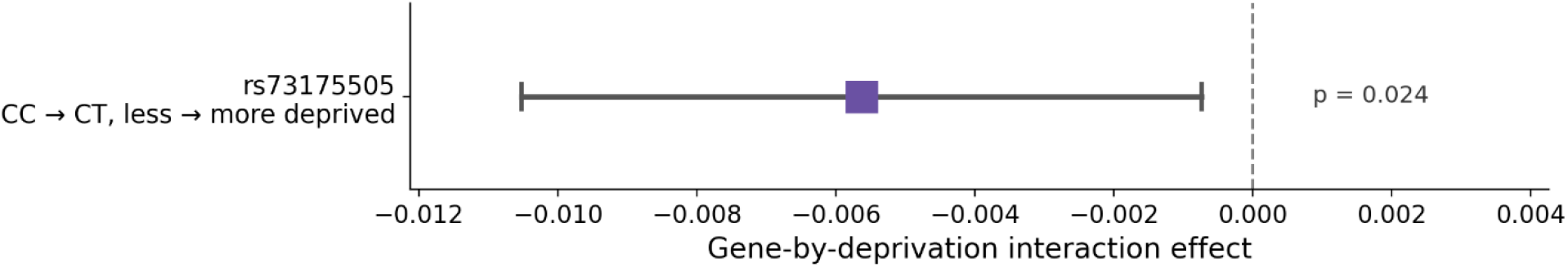
A**v**erage **interaction effect (AIE) of genotype-by-deprivation on ME/CFS risk for rs73175505 (CC→CT).** The effect is shown on the risk-difference scale for the less-deprived → more-deprived contrast, where area-level deprivation (English Indices of Multiple Deprivation) was dichotomised at the median where less-deprived falls below the median deprivation in the UKB, and more-deprived falls above the median. The bar shows the 95% confidence interval. The interaction is nominally significant (p = 0.024) but does not survive correction for multiple testing.

**Supplementary Figure 14.**
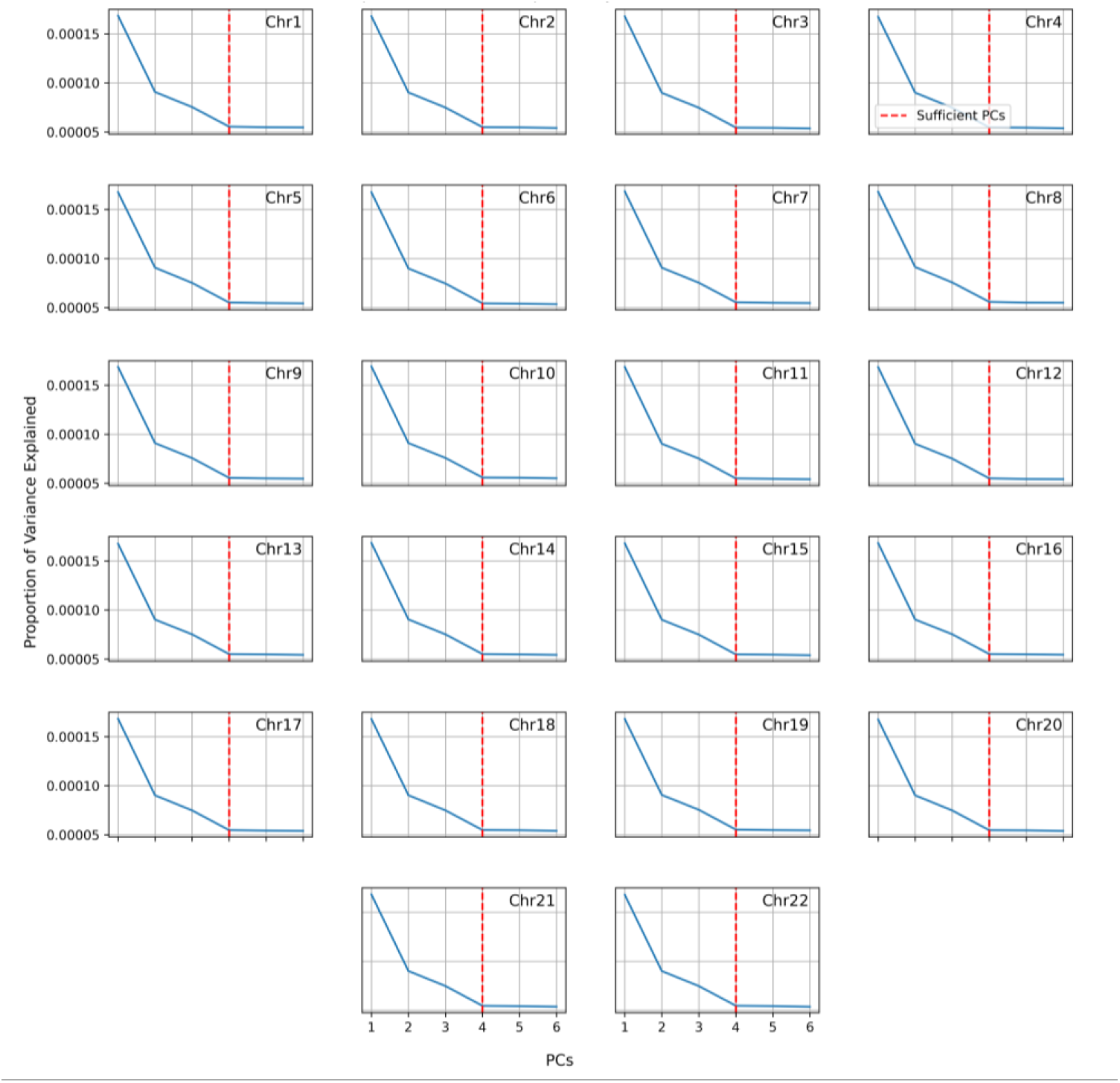
Scree plots displaying the proportion of variance explained with each Principal Component (PC) in the UKB1 population using the LOCO scheme. The label for each plot denotes the chromosome excluded from the analysis. These show that four PCs are adequate for capturing population stratification in this population. In our study we selected the set of the first six PCs, which is conservative and sufficient for the analysis.

**Supplementary Figure 15.**
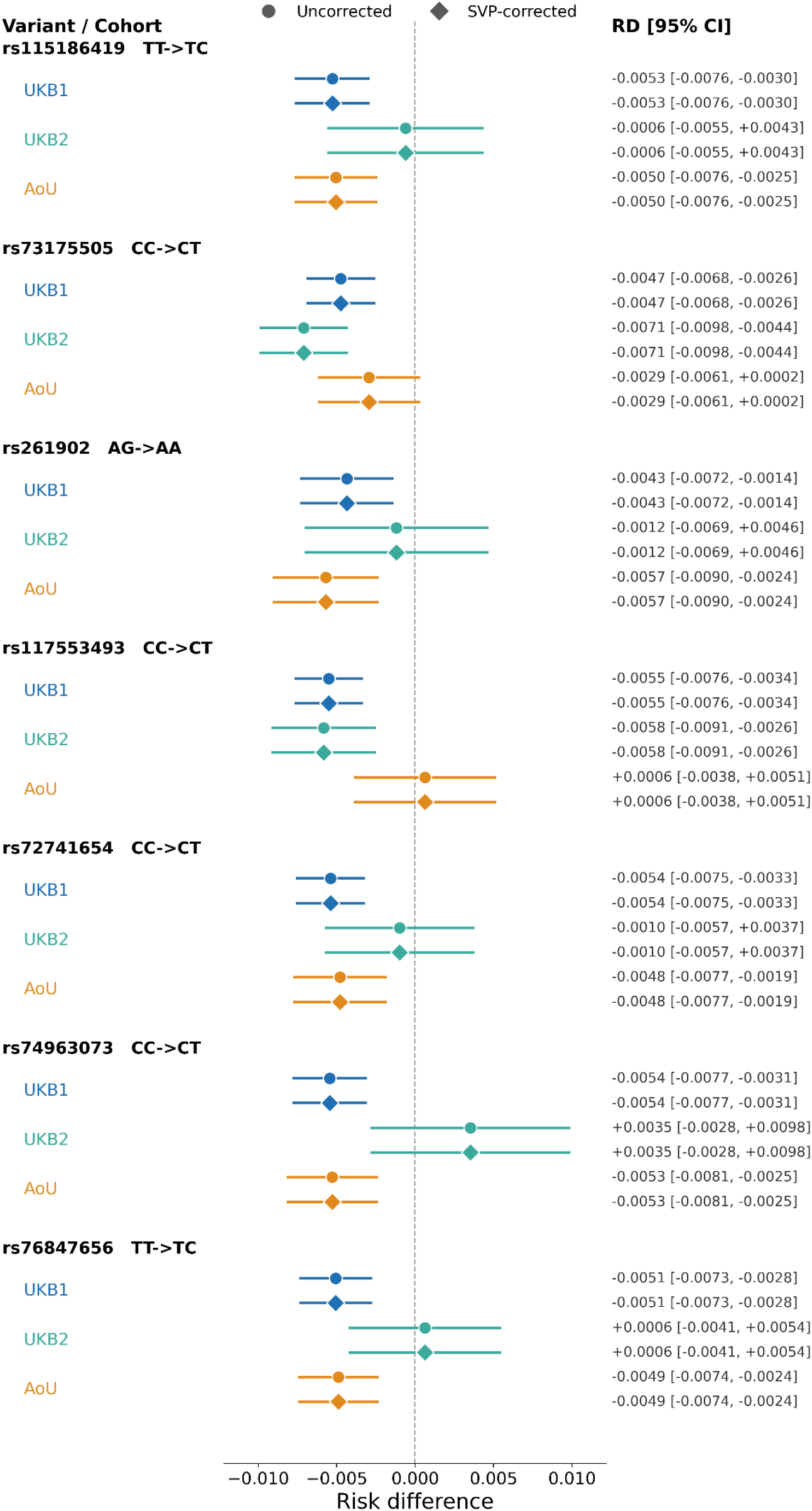
Effect estimates at the seven replicated loci are unchanged after correcting for population dependence with a Sieve Plateau Variance (SVP) estimator. The TarGene software can account for weak population dependence among participants in a study *using a class of Sieve Variance Plateau (SVP) estimators. However, this technique is computationally intensive and thus recommended in use for the estimation of genetic effects of interest in post-analyses. Here, we show the population dependence on UKB1 to be inconsequential at our replicated loci in terms of inference. Across all loci and cohorts, the SVP- corrected confidence intervals are near identical to the original estimates, indicating that accounting for population dependence does not materially alter the variance at these loci.*

**Supplementary Figure 16.**
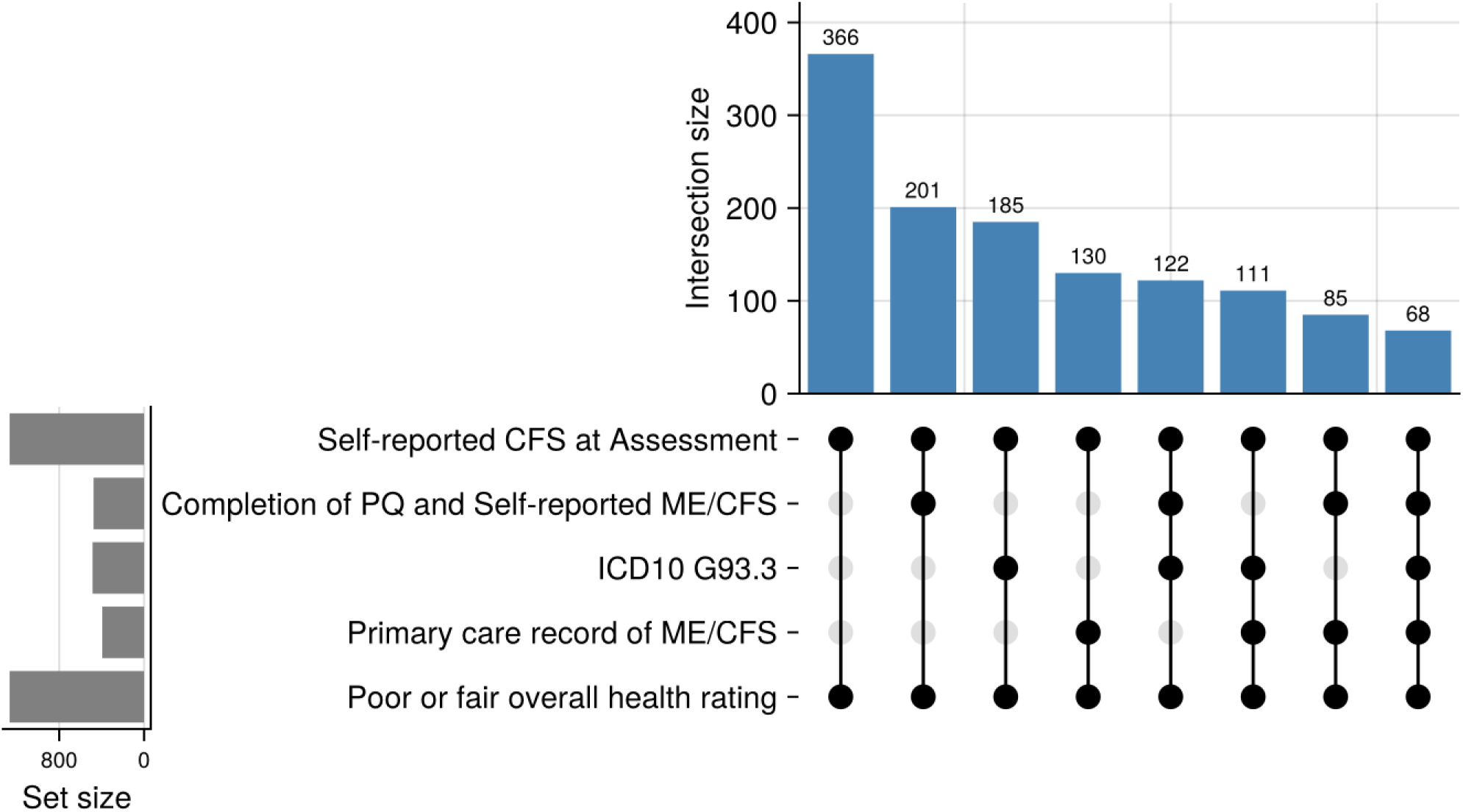
E**v**idence **for ME/CFS in UKB1.** All cases demonstrate multiple lines of evidence in affirming ME/CFS status. In UKB1, cases were expected to report a clinical CFS diagnosis at a UK Biobank assessment centre. The UK Biobank recruitment period spanned from 2006 to 2010, when cases would have reported a clinical chronic fatigue syndrome diagnosis. In 2019, the Experience of Pain Questionnaire was administered, and members of the UK Biobank cohort could optionally complete it and report a clinical ME/CFS diagnosis. 567 cases report CFS or ME/CFS from self-reported sources whereas the remaining 701 provide evidence from primary care or ICD10 codes under “postviral and related fatigue syndromes”.

**Supplementary Figure 17.**
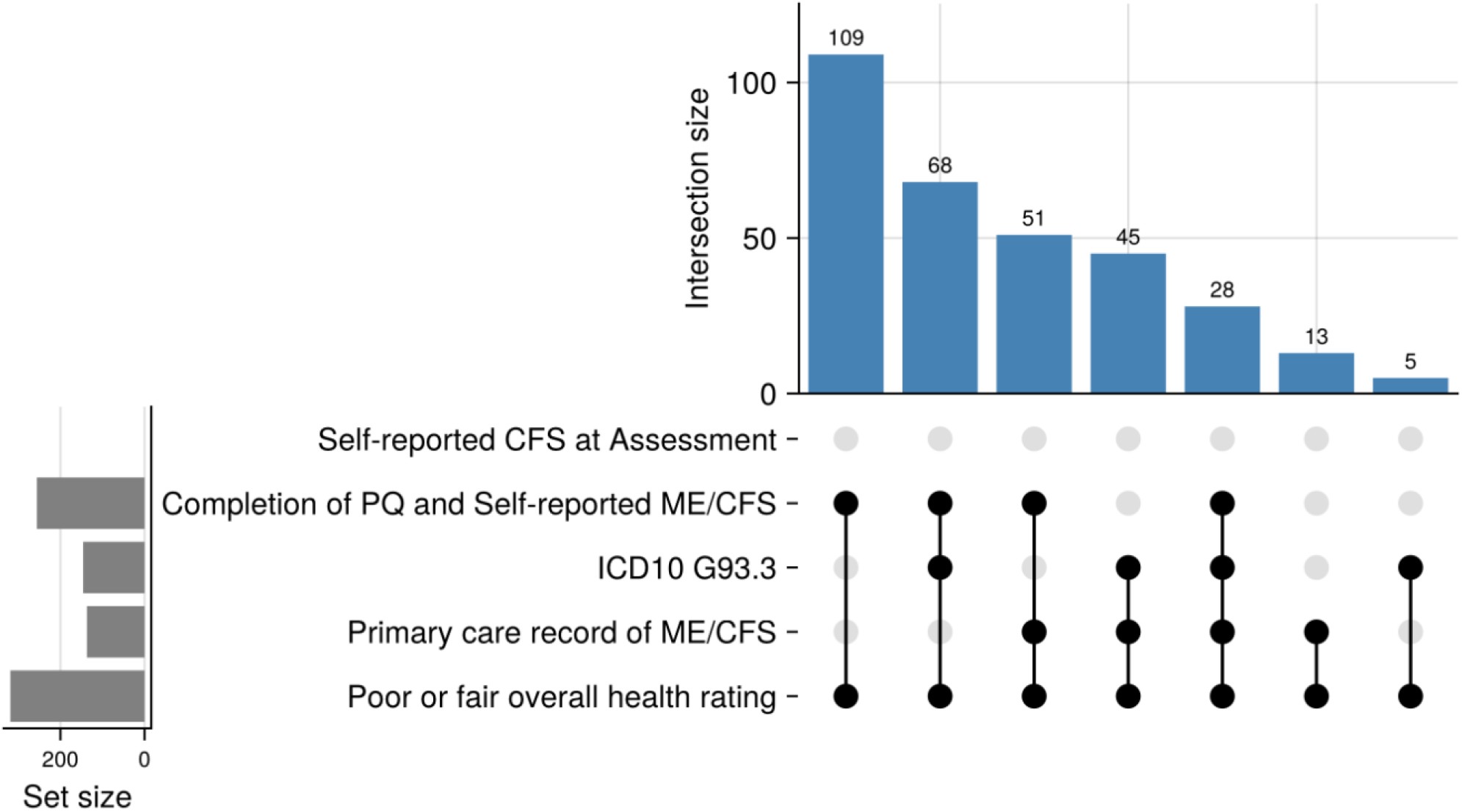
E**v**idence **for ME/CFS in UKB2.** So as to remain disjoint from the discovery cohort, UKB1, UKB2 cases were required to have not reported a clinical diagnosis of CFS at assessment but report ME/CFS through other means. Here, 109 cases reported ME/CFS through self-reporting via the pain questionnaire administered in 2019 whilst simultaneously meeting the overall health rating criterion. The remaining 210 cases reported ME/CFS via a combination of evidence consisting of at least one linkage to a primary care provider or hospital record.

**Supplementary Figure 18.**
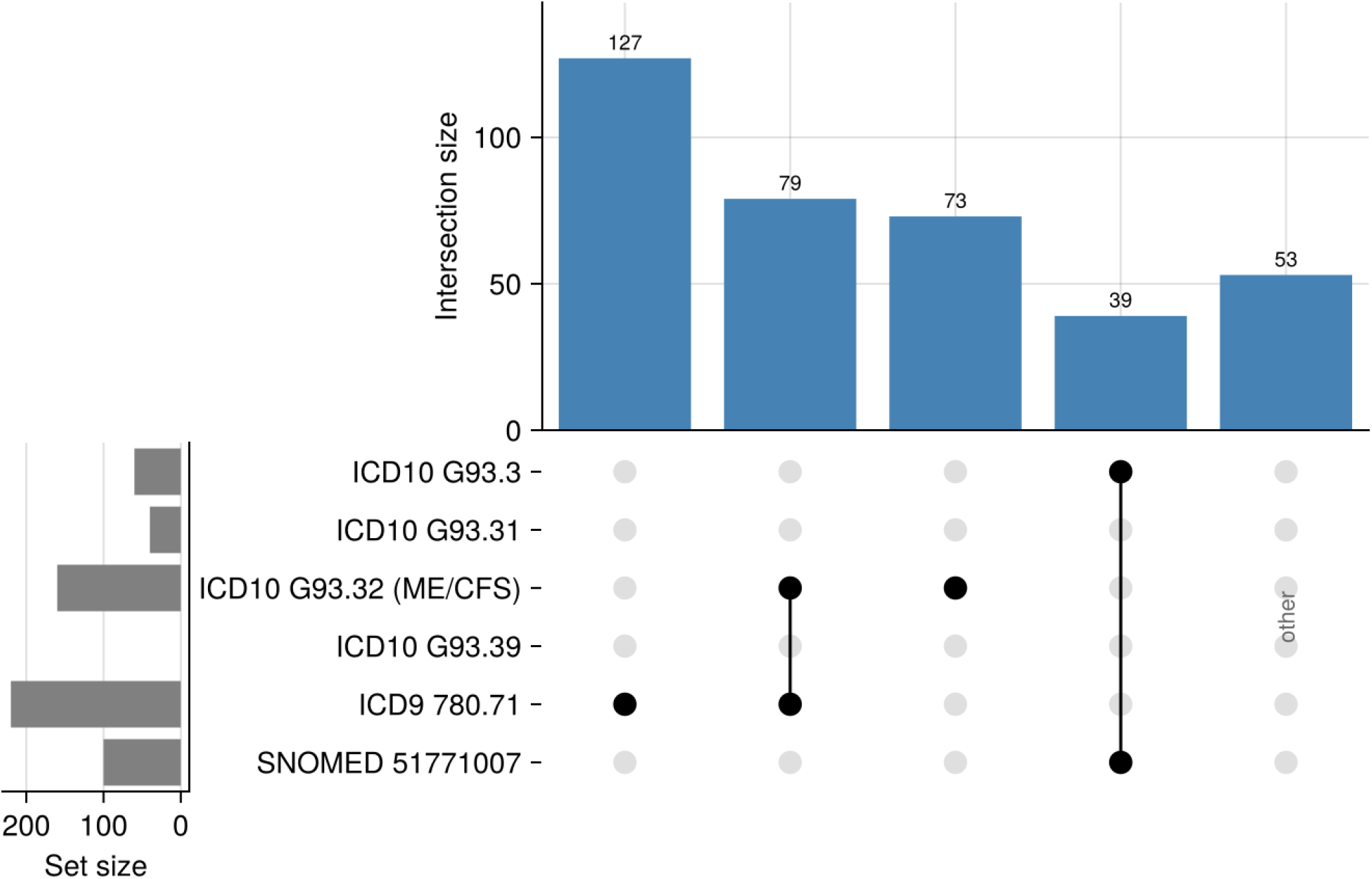
E**v**idence **for ME/CFS in AoU.** In the AoU cohort, only diagnostic labels provided by healthcare systems were used to identify participants with ME/CFS. At the request of the All of Us Research Program, we will not report participant counts of fewer than 20. *Therefore, rarer combinations of diagnostic labels will be grouped broadly as ‘other’. Additionally, because All of Us has several survey data queries relating to overall health they are omitted for readability. It should be known that all observations met the criteria for the health survey responses explicitly stated in* Table 5. The summary statistics from each of the biobank analyses can be found here with descriptions of the data on the cover sheet. ME_CFS_supplementary_tables.xlsx

**Supplementary Table 1.** Genomic risk loci identified in the analysis of imputed variants within a ±500kb window of the genotyped variants that reached genome-wide significance (p < 5×10⁻⁸) in UKB1 discovery cohort, defined by FUMA. None of these loci reached achieved replication in the disjoint cohorts (UKB2, AoU). Genomic risk loci were delineated with FUMA using the UKB release2b 10k White British reference panel for all linkage disequilibrium (LD) estimates. Candidate SNPs are all variants in the region in LD at r² ≥ 0.6 with the lead SNP. Two variants that reached genome-wide significance in the discovery analysis (rs77608528, rs115569355) were flagged as artefacts as the regions they resided in showed no linkage with other nearby variants. Positions and cytobands are reported using GRCh38. ‘Nearest gene’ is the closest protein-coding gene to the lead SNP, provided for orientation only and not implying a causal assignment.

| Locus | Lead SNP | Position (GRCh38) | Nearest gene (proximity) | Lead p-value | Candidate SNPs |
|---|---|---|---|---|---|
| 1p22.3 | rs34273700 | 1:84492521 | *RPF1* (27 kb) | 4.3 × 10<sup>−12</sup> | 3 |
| 2q11.2 | rs115349603 | 2:98543347 | *INPP4A* (intronic) | 7.2× 10<sup>−10</sup> | 30 |
| 2q13 | rs116623105 | 2:108789019 | *RANBP2* (intronic) | 1.2 × 10<sup>−8</sup> | 6 |
| 4p15.2 | rs116212207 | 4:21930291 | *KCNIP4* (intronic) | 5.3 × 10<sup>−11</sup> | 49 |
| 4q12 | rs75552905 | 4:51853931 | *DCUN1D4* (intronic) | 3.0 × 10<sup>−9</sup> | 24 |
| 9p22.3 | rs117290148 | 9:14731259 | *FREM1* (3 kb) | 3.4 × 10<sup>−9</sup> | 2 |
| 9q22.1 | rs4271043 | 9:88729856 | *NXNL2* (145 kb) | 6.7 × 10<sup>−9</sup> | 2 |
| 9q31.3 | rs117742180 | 9:110780791 | *MUSK* (intronic) | 2.6 × 10<sup>−8</sup> | 2 |
| 11q23.3 | rs7914 | 11:119309606 | *MCAM* (intronic) | 3.5 × 10<sup>−8</sup> | 2 |
| 13q32.3 | rs9585273 | 13:99907535 | *CLYBL* (intronic) | 2.0 × 10<sup>−9</sup> | 11 |
| 18q21.2 | rs140945593 | 18:54044663 | *MBD2* (106 kb) | 1.1 × 10<sup>−11</sup> | 3 |
| 18q22.1 | rs4525595 | 18:64473031 | *SERPINB8* (453 kb) | 2.9 × 10<sup>−9</sup> | 4 |

